# Dietary Modulation of The Human Small Intestine Metabolome and Microbiome

**DOI:** 10.1101/2025.11.25.25340785

**Authors:** Nadia Fernandes, Mingzhu Cai, Frederick J Warren, Anthony Duncan, Hannah Harris, Jose Ivan Serrano Contreras, Katarzyna Sidorczuk, Andres Bernal, Andres Castillo, Katerina Petropoulou, Dominic Blunt, Natalia Perez-Moral, Isabel Garcia-Perez, Cathrina Edwards, Falk Hildebrand, Elaine Holmes, Julien Wist, Gary Frost

**Author notes:** These authors contributed equally to this work (Nadia Fernandes, Mingzhu Cai, Frederick J Warren). These authors contributed equally to the generation of novel tools, analysis and construction of the manuscript (Isabel Garcia-Perez, Cathrina Edwards, Falk Hildebrand, Elaine Holmes, Julien Wist, Gary Frost).

## Abstract

The human gastrointestinal tract (GIT) is central to human physiology, yet our understanding of the metabolite, microbial, and hormonal spatiotemporal dynamics in humans is limited.

Here, we employ naso-enteral intubation to generate a spatiotemporally resolved, multimodal dataset spanning the oral cavity (salivary fluid sampling), stomach, duodenum, and ileum of ten healthy participants. In a randomised trial, participants consumed legume-based meals prepared with varying degrees of structural deconstruction, enabling investigation of how food structure shapes digestive and microbial processes along the small intestine. We find that metabolite profiles and microbial communities exhibit rapid spatial reorganisation, with evidence of transmission of oral bacterial strains to the ileum. The structural properties of food have a major impact on the metabolites and dominant microbe spatiotemporal trajectories, in the GIT thereby influencing the release of gut hormones (GLP-1 and PYY).

These findings reveal greater small intestinal metabolic and microbial plasticity during food ingestion than previously thought and suggest an underappreciated route for oral bacteria, including potential pathogens and pathobionts, to reach the ileum via the food bolus and impact metabolism. Our unique dataset allows us to explore the interplay between local and systemic metabolism, microbiota, and host physiology, with implications for metabolic health.

ISRCTN registration: ISRCTN18097249.

## INTRODUCTION

The gastrointestinal tract (GIT) is an interconnected spatially resolved organ with unidirectional flow, where different anatomical regions communicate with each other. The primary function of the GIT is the digestion of food and the absorption of nutrients, while maintaining a barrier against harmful molecules and bacteria, with the major food digestion occurring in the upper intestine. Furthermore, the GIT has a major role in fundamental homeostatic processes such as immunity, appetite regulation and glucose homeostasis^1,2^. To fulfil these varied functions, both food and the resident microbiome come together to create a complex and dynamic environment, regulated through an extensive sensing system in the GIT.

Spatially resolved areas of the GIT play specific roles not only in digestion but in the homeostatic signals they create, with over 20 hormones being secreted by endocrine cells in epithelium ^3^. For example, gastric inhibitory polypeptide (GIP) is mainly released from enteroendocrine K cells, found mostly in the duodenum, whereas glucagon like peptide-1 (GLP-1) and peptide tyrosine tyrosine (PYY), are released by L enteroendocrine cells prevalent in the ileum, colon and rectum ^4^. The product of food digestion by either mammalian enzymes or microbial action is a complex array of nutrients and metabolites which are sensed through enteroendocrine cell receptors as well as transporter networks on the epithelial surface across the GIT ^5^.

Despite its importance, the microbiome and metabolic processes located in the small intestine are far less explored than those in the distal gut, due to the inherent sample collection challenges in the former ^2,6^. 16S rRNA barcoding studies using capsule sampling ^7^ and aspirates ^8^ reveal a small intestinal microbiome dominated by *Streptococcus, Veillonella, Fusobacterium, Prevotella and Haemophilus*, with spatially variable compositions across the ileum, reflecting differences in the metabolic, pH and oxygen environments of these regions ^6^. Evidence supports the interconnectivity of microbial communities across the distinct regions of the gastrointestinal tract, with strain-level transmission observed between the oral cavity and faecal microbiome. This suggests that the oral-gut axis is more permeable than previously assumed, allowing microbial translocation along the GIT despite the physiological and immunological barriers imposed by gastric acidity, digestive enzymes, bile salts, mucosal defences, and immune surveillance, which collectively maintain compartmentalization between the oral and intestinal microbiomes ^9^. However, the prevalence and temporal dynamics of oral microbial strains in the ileum are unknown, as well as their function.

Since no studies to date have resolved the spatiotemporal metabolic, nor microbial trajectories with respect to the cellular structure of food, we developed a methodology to assess these in high temporal resolution across the oral, gastric, duodenal and ileal tract. We investigate how the molecular environment, resulting from different food preparations, impacts the microbiome in the ileum and monitor the consequent hormone release. We show that food structure strongly influences where and when nutrients and metabolites appear along the gastrointestinal tract, with broken cell meals leading to early duodenal glucose and maltose peak concentrations in the GIT fluid, while intact structures prolong nutrient delivery to the ileum. Our results reveal a marked increase in oral-derived bacterial strains in the ileum postprandially, particularly after the arrival of intact single cell or cell clusters chickpea meals. These microbial shifts are accompanied by changes in local metabolite profiles and are associated with differential secretion patterns of GLP-1 and PYY. This study provides a spatiotemporally resolved map of human digestion, showing how the food matrix modulates microbiota dynamics and enteroendocrine signalling in the small intestine.

## RESULTS

We aim to study the dynamic of spatial trajectories of metabolites and microbes across the human upper intestinal tract (oral cavity to the ileum) of 10 healthy individuals. This aim is broken down into two parts: firstly, we develop a spatiotemporal map of postprandial metabolites across the upper GIT, using an adapted naso-enteric sampling tube, to understand the impact of the cellular structure of food on molecular profiles. Secondly, we integrate the ileal metabolome and microbiome profiles to elucidate the host-microbiome interplay in processing food in the upper gut. To assess the impact of meal preparation on metabolite profiles in the gastrointestinal tract, we compared three chickpea-based meals with distinct structural integrity: broken cell (BC), clustered cells (CC) and single cells (SC). We use a panel of 35 key quantified metabolites of food digestion and microbial metabolism representing a portfolio of carbohydrate (glucose, maltose, sucrose, ciceritol, stachyose, raffinose and trigonelline), protein digestion (alanine, valine, tryptophan, glutamine, methionine, aspartate, asparagine, tyrosine, phenylalanine, histidine), bile acids (TCDCA, GCDCA, TDCA, GDCA,TCA and GCA), as well as short chain fatty acids (SCFAs) (butyrate, propionate, acetate, fumarate and formate) and key hormones. The methodology is summarised in Figure 1a and details pertaining to the study volunteers and analytical methods can be found in the “extended methods” section.

**Figure 1:**
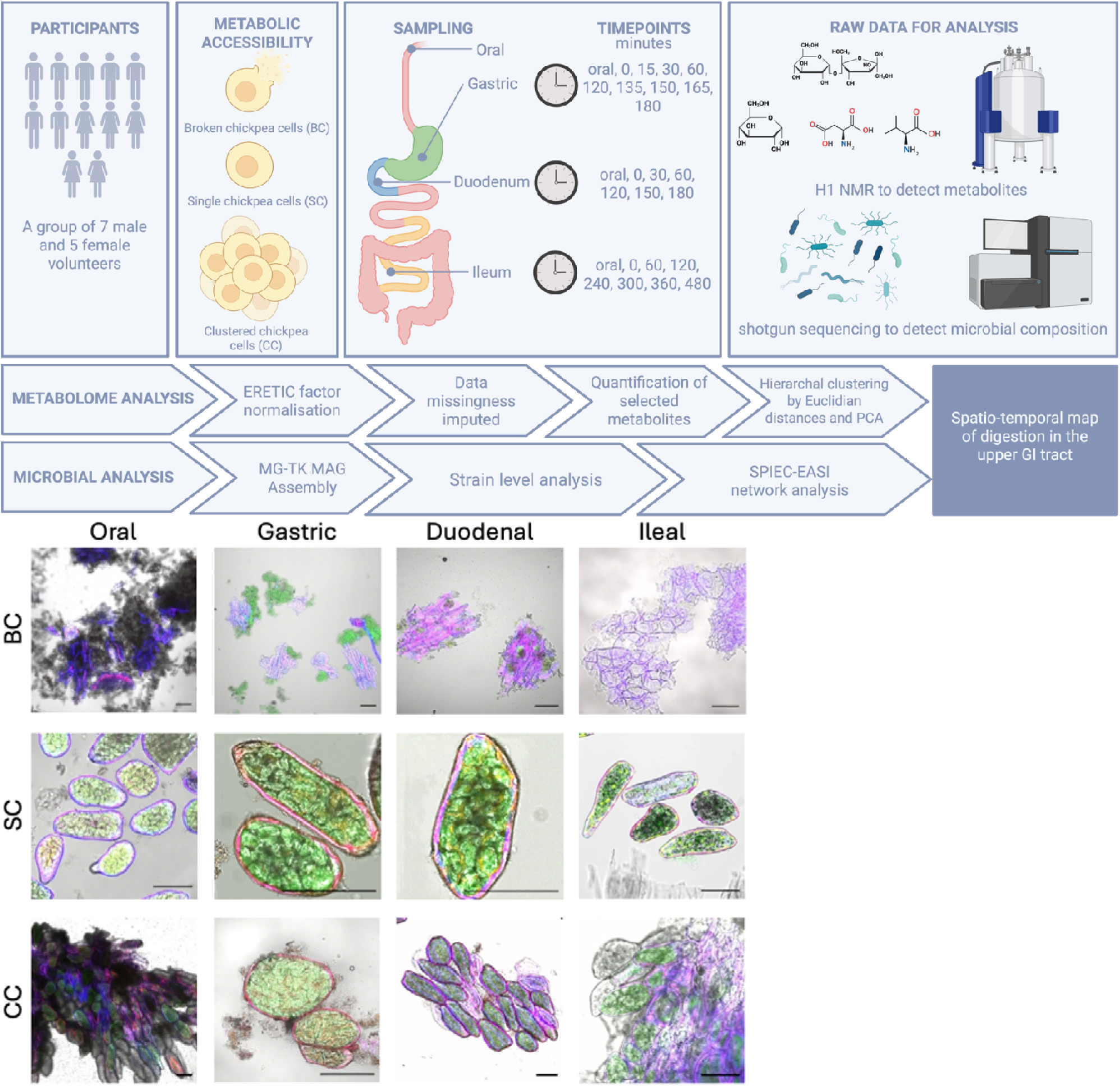
Infographic demonstrating experimental design. **(a)** Twelve participants (ten for oral, gastric, and duodenal datasets, eight for ileal dataset) were given chickpea diets in 3 different food structure matrices: broken cells (diet D0), single cells (diet D1) and cell clusters (diet D2). Naso-enteral feeding tubes were used to obtain fasted samples as well as gastric, duodenal and ileal samples at varied timepoint intervals. These samples were then processed using ^1^H-NMR and shotgun sequencing. Concentrations from NMR spectra were measured using area under the curve and data missingness was imputed using missforest R package. Euclidean distances between samples were used for principal component analysis (PCA) and hierarchical clustering. Additionally, absolute concentrations and delta concentrations were used to create 3D graphs of metabolic profiles across all three diets. **(b)** Representative confocal microscopy images of digesta aspirates collected from the mouth stomach, duodenum, and ileum of participants following consumption of a meal containing broken individual, or clustered chickpea cells. Staining highlights cell wall components (blue), protein (red), and starch (green). Scale bar = 100[sμm.

### Metabolites as endogenous markers of bolus transit

Oligosaccharides, including stachyose, raffinose, and ciceritol are synthesised in the plant cytosol ^10^. Also, trigonelline a product of niacin metabolism and a biomarker of legume intake. As these compounds are not digested or adsorbed in the small intestine, we use them as markers of intestinal flow from the oral cavity to the ileal region (Figure 2d, Extended Figures 1 and 6). The summed stachyose + raffinose (SR) concentrations relative to the gastric baseline peaked in the stomach within 15-30 minutes postprandially, within 30-60 minutes in the duodenum (relative to local baseline), and at 60 minutes in the ileum (Figure 2d). However, the timing of the peak concentration of SR varied across participants (Extended Figures 3,4,5). Approximately half of the participants exhibited a peak at 60 minutes relative to the ileal baseline, while the remainder showed later maxima (Extended Figure 5). Overall, the largest SR concentrations fold change was observed in the ileum. Trigonelline followed a similar pattern, with highest concentrations at 15-30 minutes in the stomach and 30-60 minutes in the duodenum (Extended Figure 6). In the ileum, trigonelline levels varied considerably between individuals. Ciceritol also peaked at 15-30 minutes in the stomach and at 30-60 minutes in the duodenum (Extended Figure 6), with substantial inter-individual variability in the ileum.

**Figure 2:**
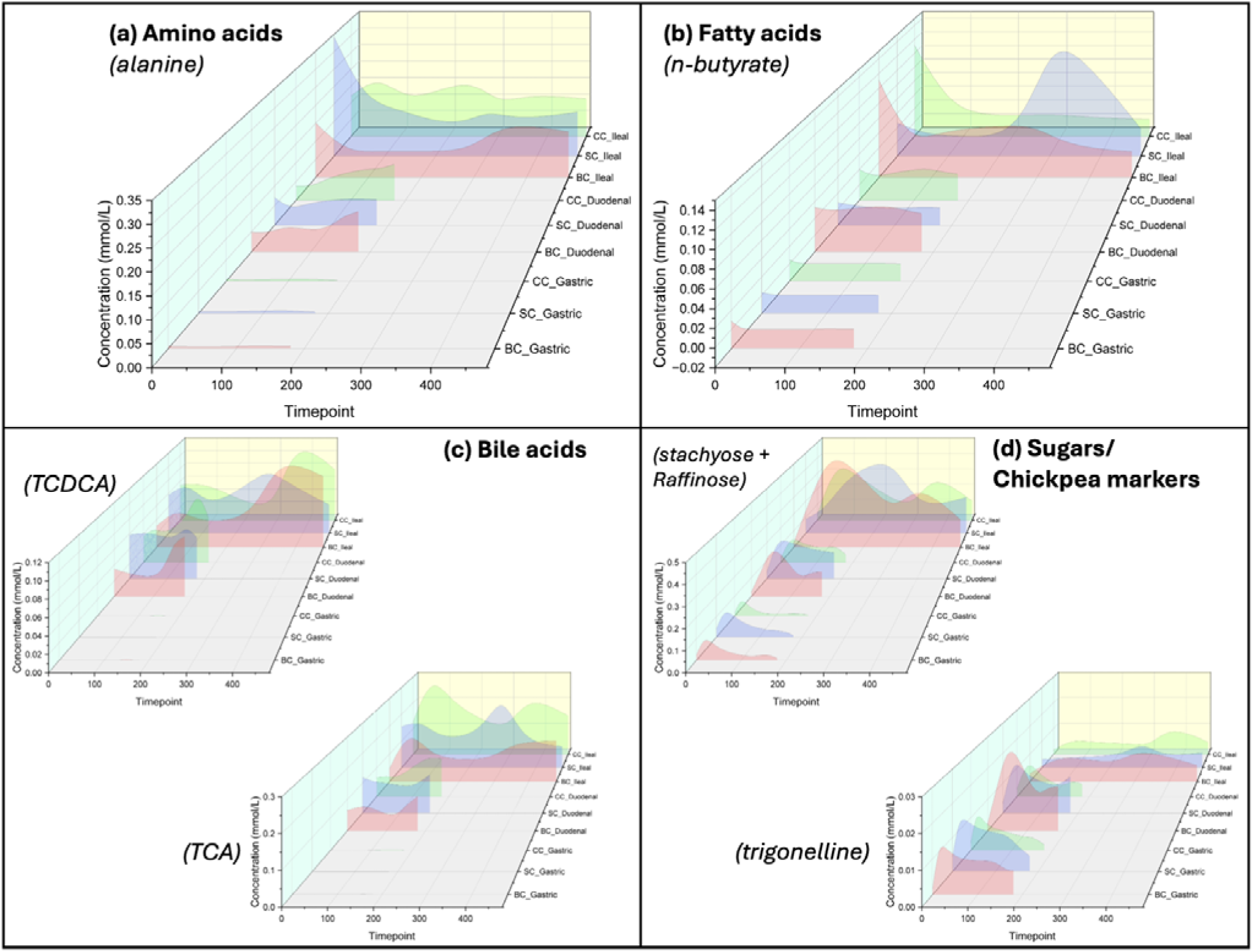
3D graphs tracking of metabolite concentrations across 3 different diets along the GI-tract. Concentrations of **(a)**amino acids, **(b)**fatty acids, **(c)**bile acids and **(d)**sugars/ chickpea markers show where Diet BC (pink) corresponds to broken cells, diet SC (blue) corresponds to single cells and diet CC (green) corresponds to cell clusters. Z-axis shows samples in the following order: three gastric samples, three duodenal samples and ileal samples. Refer to Extended Figure 1 for 3D graphs of all measured metabolites.

### Food structure influences oral-phase metabolic environment

The most abundant metabolite identified in the oral cavity prior to the bolus being swallowed was β-Maltose (10mM-50mM, Extended Figure 6), the dominant product of starch digestion, which showed a two-fold increase in BC compared to CC and SC after ingestion (∼35mM vs. 17mM, respectively). The absence of intact cell walls in BC enhanced the susceptibility of starch to salivary amylase, leading to increased maltose release during the oral phase in this diet compared with SC and CC ^11^, consistent with findings that intact chickpea cell walls limit starch bioaccessibility by slowing starch gelatinisation and restricting α-amylase access^12^.

Similarly, amino acids such as valine, tryptophan, and isoleucine were present at comparable levels in salivary fluid following ingestion of BC and SC but lower after ingestion of CC. Alanine was highest in BC and the lowest in CC (Extended Figure 6). This pattern aligns with the known role of cell wall structure in modulating nutrient bio-accessibility. The lower salivary concentrations of amino acids following CC ingestion likely reflect the reduced immediate release of intracellular solutes during oral processing due to greater structural integrity.

Oligosaccharides (SR, trigonelline, and ciceritol; Extended Figure 6) were detected at lower levels in saliva after ingestion of CC compared to BC and SC. In the oral phase SCFAs were 8-10 times lower than in the ileum (i.e., ∼0.05mM for propionate, and ∼0.5mM for acetate, Extended Figures 1 and 6). Oral acetate and propionate were lowest in BC compared to SC and CC, possibly reflecting the longer chewing time required for intact structures resulting in greater molecular release.

### Food structure does not alter gastric emptying rates

Markers of meal flow, such as trigonelline and SR (Figure 2d, Extended Figure 7), did not show significant differences in retention time across food structures (appearing in the duodenum within 30 minutes), suggesting a comparable rate of gastric emptying among the experimental meals. Previous studies have reported that particle size or microscale structure can delay gastric emptying and thus reduce glycaemic response ^13^; however, our observations at the microstructure level suggest minimal effect. Most metabolites exhibited a notable decline in concentration as the food bolus transitioned from the oral to the gastric phase. For instance, the β-maltose concentration decreased from ∼35 mM (oral) to ∼3 mM (gastric) in BC meals (Extended Figure 1 and Extended Figure 6), likely reflecting dilution by gastric secretions, a mixture of mainly water and hydrochloric acid, consistent with MRI studies showing ∼50% intragastric volume increase within 40 minutes postprandially ^14^. To control for potential confounding effects of fluid intake, volunteers were given a fixed volume of water with the chickpea meals during Visits 1 (oral) and 2 (gastric and duodenal), and no additional drinking was allowed during the sampling period. In Visit 3 (ileum), drinking was permitted but controlled as needed due to the extended 8-hour sampling protocol.

### Food structure modulates nutrient release in the gastric phase

Differences in carbohydrate concentration between food structures observed in the oral phase were maintained in the gastric phase (Extended Figure 6): β-Maltose, the marker of carbohydrate digestion, reached its peak in the stomach 15 minutes after the meal entry, with higher levels observed in BC compared to SC and CC. SR and ciceritol levels remained comparable in BC and SC but were noticeably lower in CC. Previous studies found that salivary amylase can remain active in the stomach when protected by a viscous bolus ^15^. However, laboratory studies of cellular chickpea porridge meals suggest that these do not form a cohesive bolus, and that there is limited continuation of starch digestion by salivary amylase in the stomach ^12^. Our data also suggest that the compact structure of CC further resists mechanical breakdown in the stomach, resulting in reduced nutrient release relative to BC and SC. Amino acids and SCFAs too were detected at lower concentrations in the stomach (0.01-0.05mM), with a prolonged release phase initiated after 120 minutes, particularly for valine, asparagine, histidine, and SCFAs, not differing substantially between intact and broken food structures (Figure 2a, 2b and Extended Figure 1).

### Nutrient release and absorption in the duodenum

The duodenum is the first segment of the small intestine for food digestion and nutrient release and absorption. SR (Figure 2d) increased in the duodenum from 30 min, peaked around 60 min and gradually returned to baseline over 180 min, indicating the timeframe of food retention in the duodenum. Higher levels of SR were observed in BC compared to CC postprandially at 60 min, as observed in the gastric region (Extended Figure 6). Starch digestion products, β-maltose and glucose (Extended Figures 1 and 6), were the dominant metabolites of chickpea digestion in duodenal fluids. Differences between food structures were most pronounced at 60 minutes, where BC resulted in 1 mM β-glucose and 5 mM β-maltose, representing a 3–4-fold increase compared to SC and CC, in alignment with the expected bioavailability for all 3 preparations. Amino acid concentrations increased from the gastric to the duodenal phase, particularly after 120 minutes postprandial (∼0.05–0.1 mM). Amino acids including valine, alanine, glutamine, and histidine appeared higher in SC and CC than in BC (Figure 2a and Extended Figure 1), confirming previous observations ^1^. Duodenal bile acid profiles were modulated by food structure, with taurocholic acid (TCA), and glycocholic acid (GCA), exhibiting higher maxima in SC than in BC during 30-120 minutes (Figure 2c and Extended Figures 1 and 6). Other bile acids, including glycochenodeoxycholic acid (GCDCA), taurodeoxycholic acid (TDCA), glycodeoxycholic acid (GDCA) were comparable across meals. SCFAs were more abundant in the duodenum compared to the gastric phase (Extended Figure 1), with significant differences observed between food structures for butyrate (Extended Figure 6) that shows increasing levels for BC and CC during 120-180 minutes.

### Microbial metabolism and nutrient dynamics in the ileum

The ileum is the final segment of the small intestine and plays a key role in integrating digestive, microbial, and hormonal signals. We observed that the stachyose + raffinose (SR) marker of intestinal flow peaked in ileal samples around ∼60 min post-ingestion (Figure 2d). This is consistent with findings by Dagbasi et al. (2024) ^16^, who showed that food-derived metabolite arrival in the ileum from ∼60 min contributes to PYY secretion through L-cell activation in the distal gut ^16^.

We next examined how food structure influenced the arrival of digestible nutrients to the ileum. Meals composed of single (SC), or clustered (CC) chickpea cells led to higher concentrations of β-glucose and β-maltose in the ileum compared to broken-cell (BC) meals (Extended Figure 1, Extended Figure 6). Again, this reflects the reduced starch digestibility in the proximal gut due to intact cell walls in SC and CC, enabling more digestible carbohydrate to reach distal regions. Similarly, amino acid concentrations were generally higher in the ileum than in the duodenum, with SC samples showing particularly elevated levels of valine, glutamine, and methionine, tyrosine, phenylalanine, tryptophan, histidine at 120 min.

Short-chain fatty acids (SCFAs) in the ileum were highest in the fasted state and dropped sharply after food intake (Figure 2b, Extended Figure 1). From 240 min onwards, SCFAs gradually increased, more specifically in the SC group, where we observed elevated n-butyrate, propionate, and acetate. Since these profiles reflect delayed fermentation and downstream microbial activity, substantial inter-individual variation was observed (Extended Figure 5). Formate, which was consistently higher in the ileum than in gastric or duodenal samples, showed only minimal postprandial fluctuation, suggesting it may represent a highly regulated microbial or host-derived metabolite in this region.

We also observed spatial and structural differences in bile acid dynamics across the gut. Total bile acid concentrations increased progressively from the duodenum to the ileum, peaking between 300–360 min (Figure 2c, Extended Figure 1). Among primary bile acids, CC meals yielded higher TCA and GCA concentrations at 60 and 120 min. In contrast, TCDCA and GCDCA were elevated in SC compared to CC and BC at later timepoints. For secondary bile acids, CC samples exhibited higher levels of TDCA than SC during late-phase ileal sampling (Extended Figure 6). These differences likely reflect both the timing of substrate delivery and the extent of microbial activity (deconjugation), modulated by food microstructure.

As expected, substantial inter-individual variability was observed across all metabolites, in baseline levels, peak timing, and postprandial magnitudes (Extended Figures 3-7). Variability was more pronounced in microbiota-related metabolites (e.g., SCFAs) compared to food-derived metabolites (e.g., carbohydrates and amino acids). Also, ileal metabolites displayed greater variability compared to other GIT regions. This may be attributed to differences in microbial species and activities among individuals.

### Relationship between metabolite levels and gut hormone release

To investigate links between metabolites and gut hormone responses, we used hierarchical clustering analysis (HCA) (Figure 3b) to reveal associations between metabolic profiles and plasma concentrations of GIP, GLP1, and PYY (Figure 3a). For interpretability, we arbitrarily divided the postprandial window into three time-segments: 0–30 minutes (hormone induction phase), 30–120 minutes (hormone maintenance phase), and 120–480 minutes (hormone extension phase).

**Figure 3:**
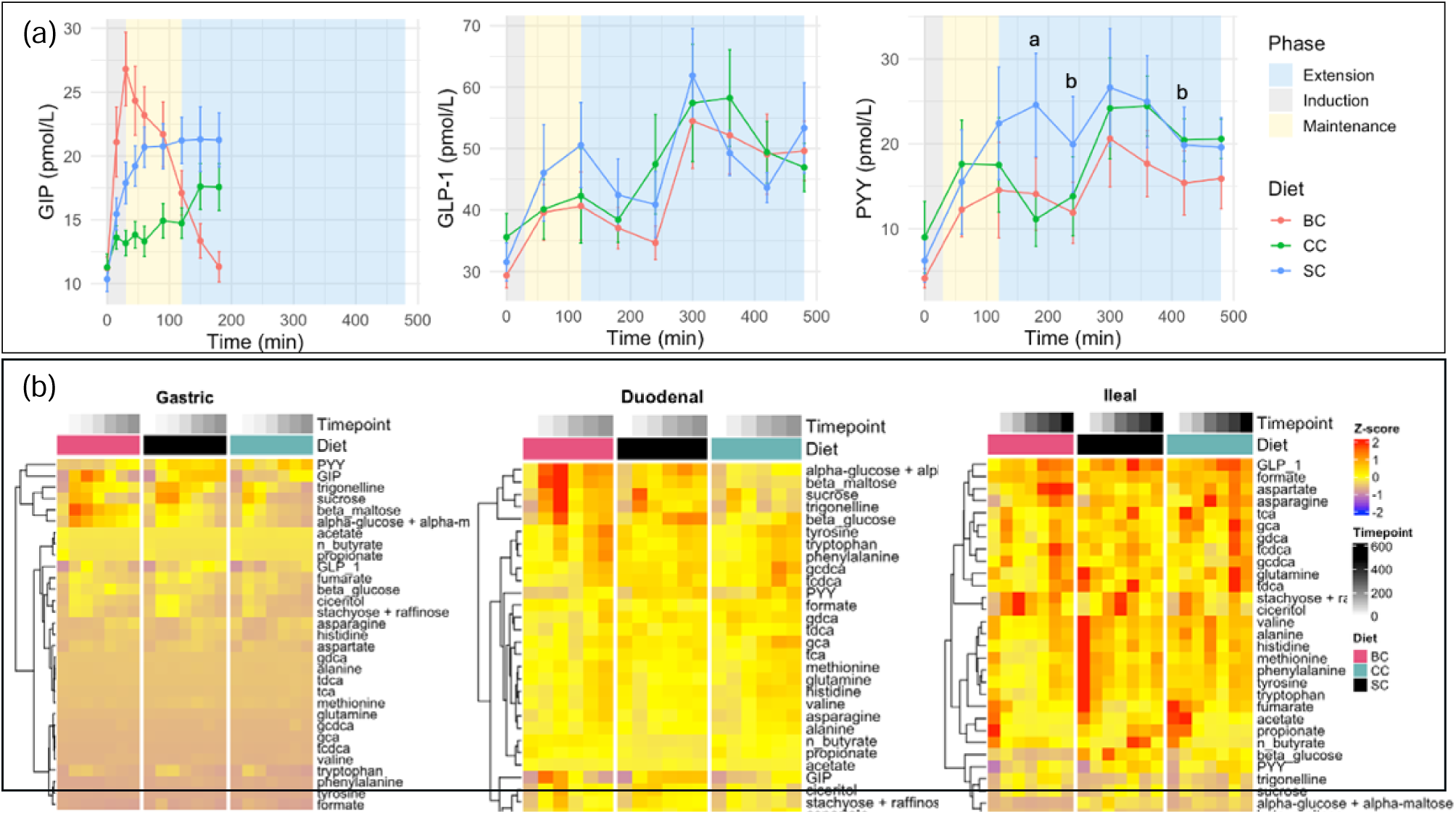
**(a)** Time-course changes in GLP-1 and PYY following the consumption of chickpea meals (blue: Intact single cells (SC), green: cell clusters (CC), red: broken cells (BC)). Data are presented as mean ± SEM. Statistical differences between groups were analyzed using repeated measure two-way ANOVA followed by Tukey’s post-hoc test (p < 0.05). Different letters (a, b, c) indicate significant differences between groups at the same time point: a – SC vs CC; b- SC vs BC. **(b)** Relative concentration of metabolites and hormones per timepoint, across meals, along the GI-tract.

We found that GIP release during the initiation and early maintenance phases was associated with the presence of maltose in the gastric and duodenal environments. Broken-cell (BC) meals, which released more maltose proximally, were linked to higher GIP levels, while clustered-cell (CC) meals, which released less maltose, showed attenuated GIP secretion.

In contrast, GLP1 and PYY exhibit more complex dynamics. Early PYY release correlates closely with GIP, suggesting a shared proximal stimulus in the initial postprandial phase. GLP1 initiation, however, is more strongly associated with carbohydrate metabolites in the stomach and duodenum than with other hormones.

Interestingly, fasting SCFAs levels, independent of the meal structure, in the ileum were positively associated with early PYY secretion across diets (Figure 3b). While chickpea-derived SR appeared in the ileum by ∼60 minutes, the timing suggests that early SCFAs-linked PYY stimulation may be influenced by baseline or proximal microbial cues. No similar association was found for GLP1 in this early phase.

During the late maintenance phase, a second rise in GLP1 and PYY levels corresponded to the arrival of diverse nutrients in the ileum. PYY was associated with a mix of carbohydrate and amino acids, while GLP1 was more strongly linked to formate, aspartate, and asparagine. Notably, bile acids are more closely linked with GLP1 than with PYY (Figure 3b), suggesting that GLP1 secretion may be more responsive to bile acid signaling in the ileal environment.

Finally, food structure had a marked effect on interactions between hormones and metabolites. BC meals enhanced carbohydrate exposure in the stomach and duodenum and were associated with stronger GIP release (Figure 3a). In contrast, SC meals were more effective in sustaining GLP1 and PYY levels, likely due to delayed nutrient release into the ileum (Figure 3a). At later timepoints (∼420 min), CC meals resulted in elevated carbohydrate and amino acid levels, coinciding with a convergence of PYY responses between SC and CC meals (Figure 3a, 3b).

### The microbiome composition of the ileum is dynamic

Analysis of the microbiome composition of the ileum during consumption of a meal demonstrates the highly dynamic nature of the microbial ecosystem. In the fasted state the microbial community of the ileum of the volunteers is dominated by the genera *Haemophilus* and *Streptococcus*, including contributions from facultative anaerobes which may be considered from oral or upper gastrointestinal tract taxa such as *Gemella, Veillonella, Peptostreptococcus, Actinomyces, Rothia* and oral *Streptococcus* species (Figure 4c) ^6^. We also note in our volunteers in the fasted state the presence of strict anaerobes generally characteristic of the colonic environment such a *Bifidobacterium, Veillonella, Bacteroides,* and *Faecalibacterium prausnetzii* (Figure 4c) ^6^. Following ingestion of a meal we observe rapid changes in the community composition of the ileal microbiome. The arrival of the food bolus in the ileum, between 60 and 120 minutes, is associated with a decrease in microbial numbers^16^, but an increase in microbial community diversity (Shannon index) for the BC and CC meals, but not for the SC meal (Figure 4a). There is a later secondary increase in microbial diversity at between 360 and 480 minutes for all three preparations, which is likely due to the arrival in the ileum of the second standard pasta-based meal given in the study and coincides with the second peak in SR (Figure 2 and Figure 4). The arrival of the meals in the ileum changes the relative abundance of several species (Figure 4c). Species such as *Rothia aeria* and *Rothia denticariosa* increase in relative abundance following consumption of a meal, while species such as *Klebsiella varicola*, *Romboustia timonensis* and *Haemophilus parainfluenzae* are observed to decrease. The species with increased relative abundance are typically found in the oral cavity, while those which decrease are more commonly considered colonic anaerobes. We hypothesise that this may be indicative of the bolus carrying oral bacteria into the ileum, replacing the resident colonic bacteria.

**Figure 4:**
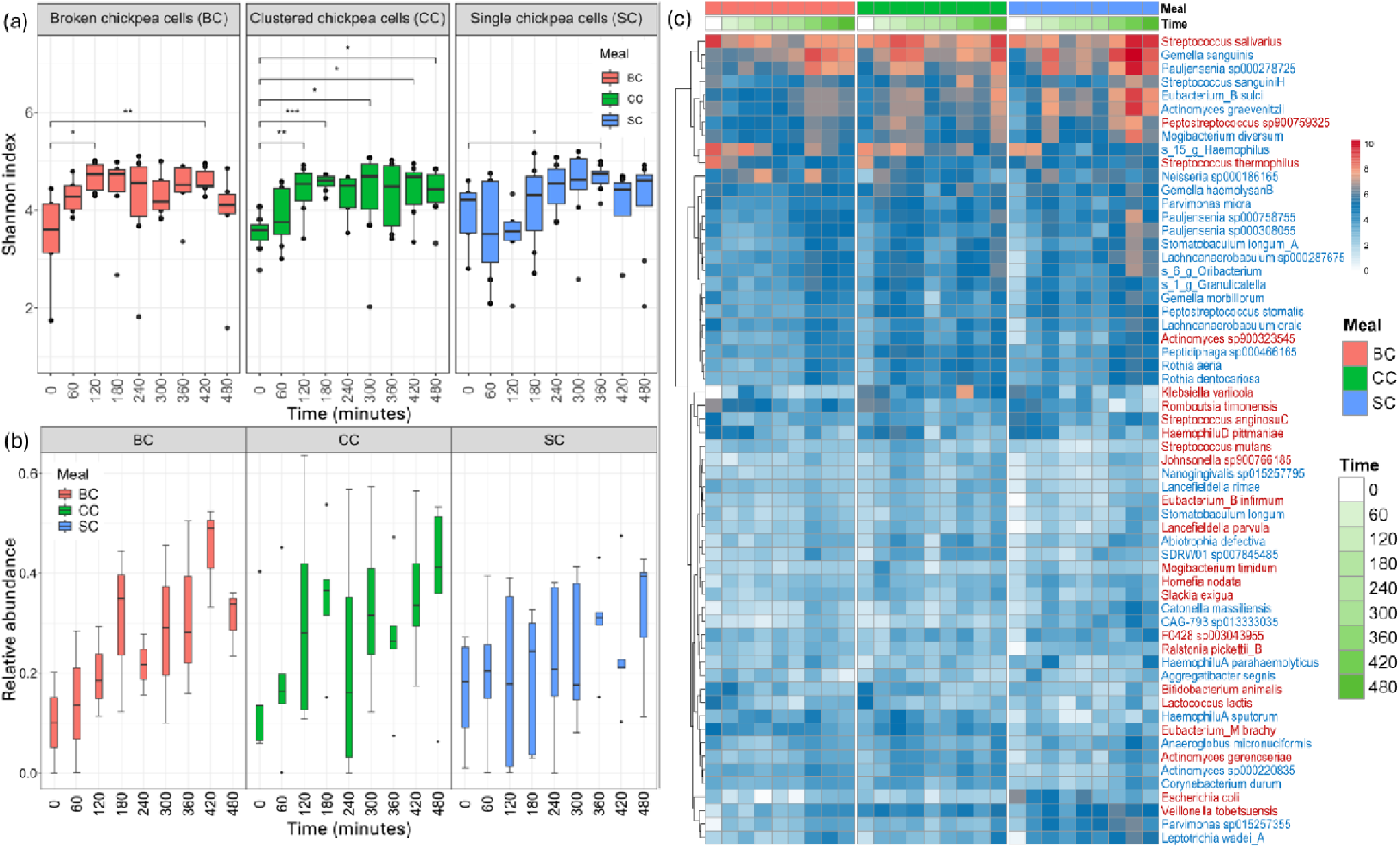
Variation in diversity and abundance of oral and ileal strains. **(a)** Alpha diversity changes as a function of time in the ileum. Shannon diversity index changes over time for meal BC, CC and SC. Statistical significance is indicated as a change relative to baseline, using a Wilcoxon test with a p-value cut-off of 0.05 for statistical significance. **(b)** Proportion of strains shared between oral and ileal compartments as a function for the total number of strains identified, for each of the meals consumed. **(c)** Relative abundance (log2 transformed) of the 50 most abundant species in the terminal ileum over time during consumption of each of the 3 diets. Species names highlighted in red did not show evidence of oral-ileal strain sharing. Species names highlighted in blue, did show evidence of oral ileal strain sharing.

### Evidence of strain sharing between oral and ileal compartments

To test the hypothesis that the oral and ileal microbial communities may mix while consuming a meal, we acquired strain-resolved metagenomes of saliva samples from five participants. Hildebrand et al. (2021) ^17^ Figure 4b provides direct evidence of strain sharing between the oral and ileal compartments. Across the 5 participants for which we had paired data, we identified 106 strains that were shared between the oral and ileal samples. To quantify the proportional contribution of strains shared between oral and ileal samples, we analysed microbial abundance in the ileum during meal ingestion (Figure 4b). We observe an increase in the proportion of oral strains in the ileal microbiome following ingestion of a meal, indicating that the arrival of the bolus in the ileum is associated with translocation of bacteria from the oral cavity. In the fasted state, around 10% of the total bacterial abundance in the ileum can be attributed to oral species. This rises with ingestion of the meals, with a first peak at around 120 minutes corresponding to the arrival of the chickpea meal in the ileum, and a second peak at 420-480 minutes following lunch to values for the as high as 50% of the total microbial community (Figure 4c). Several oral species, most notably *Gemella sanguinis* are observed to increase in relative abundance the ileum. Other highly prevalent shared species include *Mogibacterium diversum*, *Actinomyces graevnitzii* and *Streptococcus sanguinis*.

### Interactions between oral microbial strains, metabolites and gut hormones in the ileum

Co-occurrence networks constructed from the strain resolved microbiome data and metabolites at baseline, 120 and 480 minutes show a lack of interactions between metabolites and microbial species at baseline (Figure 5a-d), and most of the species observed had a low proportion of strain sharing between oral and ileal compartments. The species with a higher proportion of strain sharing are mainly clustered around *Eubacterium sulci*, which showed negative associations with other parts of the network. A distinct, tightly clustered module of colon-associated strict anaerobes centred on *Faecalibacillus intestinalis,* with no evidence of strain sharing with the oral cavity. The baseline network has a high degree of modularity, with clear clustering of species, and the modularity reduces over time. At 120 minutes the network became more diverse, with a wider distribution of nodes showing evidence of strain sharing between oral and ileal compartments. At 480 minutes the network was more tightly integrated with the majority of hubs within the clusters showing evidence of strain sharing. The cluster of colonic anaerobes centred on *F. intestinalis* observed at 120 minutes is much less tightly connected, and the abundances of the nodes are much lower in contrast to the shared nodes.

**Figure 5:**
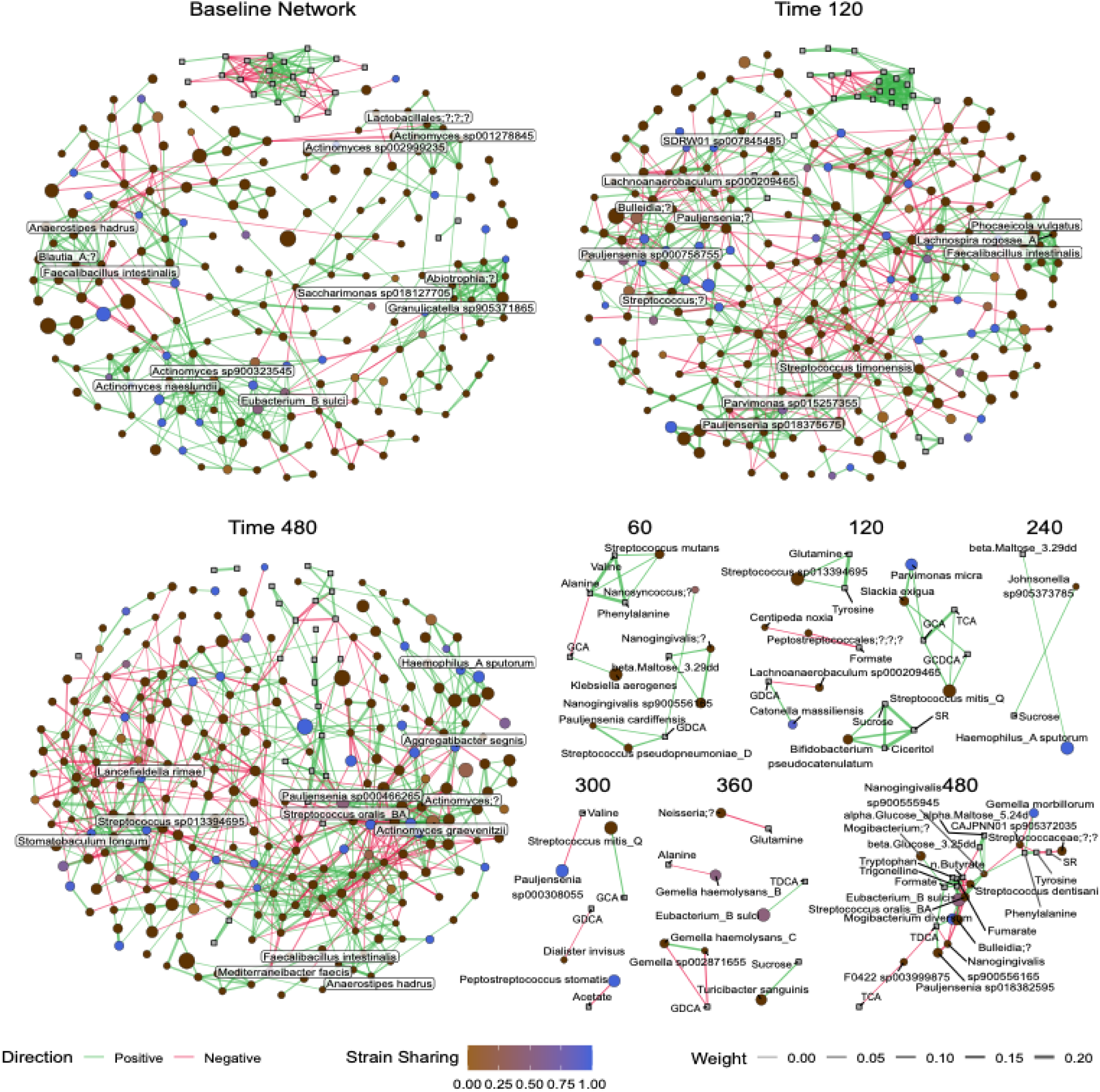
Co-occurrence analysis between ileal species and metabolites over time. Dense co-occurrence networks showing all microbial taxa detected, all interactions and all metabolites in the terminal ileum at **(a)** baseline, **(b)** 180- and **(c)** 420-min following ingestion of a meal. **(d)** The sparse networks are filtered to show only the metabolite-microbe interactions, and only for the timepoints where there are significant microbe-metabolite interactions. Nodes are colored by the degree of strain sharing between oral and ileal compartments. Nodes in red are not shared, while nodes in blue are shared between the oral and ileal compartments.

Microbe–metabolite associations were most prominent at 60–120 and 360–480 minutes, aligning with the meal arrival in the ileum (Figure 5d). These associations involved SCFAs (acetate, n-butyrate, formate), amino acids (glutamine, phenylalanine), and various bile acids. During early timepoints, associations were primarily observed with non-adherent, shared strains, suggesting that microbial metabolism of freely available substrates (e.g., sugars, amino acids) may precede visible particle degradation. This aligns with previous findings of early GLP1 release in the absence of visible microbial colonization. At later timepoints (360–480 min), associations increasingly involve strains with oral origins, including *E. sulci* and *M. diversum*, highlighting a potential role for these transmitted taxa in shaping ileal metabolite profiles.

We further investigated associations between microbiome composition in the ileum and hormone release (Extended Figure 10). Only few taxa were identified that correlated to GLP-1 release, mainly with species that did not show evidence of strain sharing such as *Abiotrophia defective* and *Pauljensenia bouchesdurhonensis*. In contrast, we observed a larger number of associations between PYY release and ileal microbiome composition. While species that were negatively associated with PYY release were colonic species with no evidence for strain sharing, many of the species positively associated with PYY release were shared with the oral cavity. There was some overlap observed between the species associated with individual metabolites and hormone release, for example *E. sulci* was associated with higher n-butyrate levels and with higher PYY release.

### Metabolite and microbial patterns in the ileum influence plasma gut hormone profile

As well as taxonomic changes over time, we also identified shifts in functional pathways within the microbiome following arrival of the bolus (Figure 6a). There are shifts in the abundance of several microbial pathways related to carbohydrate metabolism (starch, lactose and sucrose degradation, glycolysis) and SCFAs production (lactate production, acetyl-CoA to acetate and pyruvate formate lyase) over time for each of the meals.

**Figure 6:**
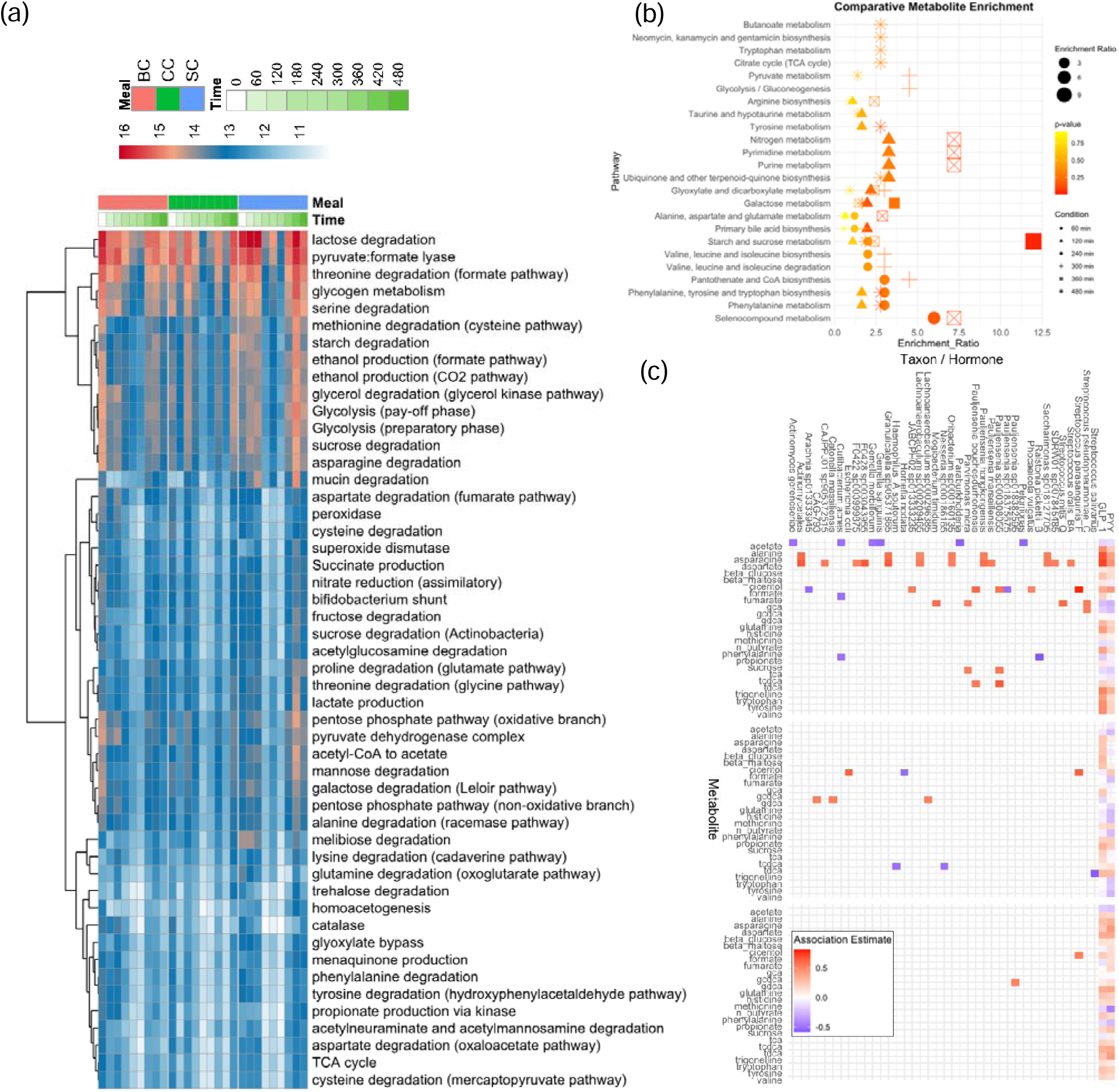
Microbiome- and metabolome-based functional analysis. **(a)** Heatmap showing KEGG pathways predicted using the microbiome data. **(b)** KEGG-based metabolic enrichment analysis across timepoints. A subset of metabolites, identified through significant microbe-metabolite associations in network analysis, was analyzed to reveal pathway-level changes at each timepoint. Metabolite enrichment analysis was performed in MetaboAnalyst using a custom background of all measured metabolites. **(c)** Highly significant taxon and hormone associations with metabolite per diet; BC (upper box), SC (middle box) and CC (lower box), derived using SEM analysis.

As a complement to this analysis, metabolite set enrichment analysis (Figure 6b) of microbiome-associated metabolites (Figure 5d) revealed temporally coordinated shifts in ileal metabolic activity following chickpea ingestion (timepoint 0). At 60 minutes, phenylalanine metabolism, aromatic amino acid biosynthesis, and pantothenate and CoA biosynthesis were enriched, indicating early microbial involvement in amino acid processing and cofactor production; selenocompound metabolism was also enriched at this time, though this may be attributable to the metabolite subset captured in the NMR panel. By 120 minutes, pathways linked to primary bile acid biosynthesis, galactose metabolism, and glyoxylate/dicarboxylate metabolism were activated, suggesting early host-microbial coordination around bile acid signalling and sugar utilization. At 240 minutes, strong enrichment of starch, sucrose, and galactose metabolism was observed shortly after the first ileal sugar peak (Figure 2), consistent with microbial engagement in carbohydrate breakdown and utilization. At 300 minutes, pantothenate and CoA biosynthesis re-emerged, accompanied by glycolysis/gluconeogenesis and pyruvate metabolism, reflecting microbial energy harvesting from residual substrates. By 360 minutes, purine, pyrimidine, and nitrogen metabolism dominated, consistent with microbial nucleotide synthesis and recycling; selenocompound metabolism was also enriched at this time, though again this may reflect the metabolite subset captured in the NMR panel. Finally, at 480 minutes, phenylalanine metabolism, aromatic amino acid biosynthesis, and tyrosine metabolism were again prominent, marking a late-phase shift toward aromatic amino acid transformation likely driven by reduced carbohydrate availability and the use of amino acids as alternative substrates and biosynthetic precursors. Collectively, these results suggest that following a chickpea meal, the ileal microbiome engages in a sequential program of amino acid, carbohydrate, and nucleotide metabolism, reflecting dynamic host-microbe co-metabolism across distinct postprandial phases.

We further modelled the associations between microbiome, metabolites and gut hormones for each of the separate meals (Figure 6c). We identified more associations between microbial species and metabolites for meal BC compared to meals SC and CC. Between all three meals we identified a consistent positive association between the abundance of the *S.parasanguinis* and the presence of the microbial metabolite formate, which was also positively associated with PYY signalling in all the meal conditions tested. *S.parasanguinis* is a well characterised formate producing oral microbe, which generates formate from sugar metabolism via heterolactic fermentation ^18^. Several other predominantly oral derived species were associated with formate for both meal BC and meal SC, including several *Pauljensenia* species. Formate was observed to increase in the ileum following arrival of a meal (Extended Figure 1), unlike the other SCFAs which rapidly reduced, suggesting that formate may be a key metabolite produced by tissue in ileum in the fed state, consistent with a predominance of oral bacteria. Conversely, only negative microbial associations were observed with acetate and propionate, which were also negatively associated with the gut hormones PYY and GLP-1. For meal BC there were also strong associations between a large number of microbial taxa and the amino acids asparagine and aspartate, which were associated with GLP-1, and to a lesser extent PYY release, although these microbial associations were not observed for meals SC and CC.

## DISCUSSION

### Our work describes for the first time the flow of metabolites in the GIT from the oral cavity to the ileum

Studies of the GIT metabolites and microbiota have relied mainly on indirect methods of assessment such as stool or animal models. Recent studies, enabled by gut intubation and sampling methodologies, examined metabolite profiles in isolated GIT segments, such as the ileum ^16^, and the duodenum ^19^. A comprehensive picture across the entire upper GIT has been lacking.

Our study addresses this gap by providing a continuous mapping of metabolite flow from the oral cavity through to the stomach, duodenum, and ileum. This offers an integrated perspective on digestion and nutrients availability across the GIT, which is crucial for understanding the nutrient sensing and the way the human GIT communicates across the GIT. Sensing of nutrients by chemosensory cells across the GIT plays a key role in transmitting food-related signals to organs and tissue that generate physiological regulatory responses. Spatially specific enteroendocrine cells (EECs) across the GIT respond to luminal nutrient stimuli and secrete gut hormones ^20^, for example, K-cells predominately located in the duodenum release GIP ^21^, while L-cells residing in the ileum and colon release GLP-1 and PYY ^22^. The distribution and flow of nutrients through distinct GIT regions influence the release patterns of these gut hormones, which regulate local digestive processes and control key physiological functions such as insulin secretion and appetite control ^22,23^.

By tracking metabolite concentrations across multiple GIT regions, **we show a major impact of food structure both on molecular environment and ileum microbiome**. Current dietary strategies to combat non-communicable disease focus on specific food or nutrient intake but overlook the role of food structure. Previous studies have shown that processed and unprocessed diets, matched for macronutrient and energy density, have different effects on appetite-regulating hormones and body weight ^24^. One proposed mechanism is that the loss of cellular structure in the processed diet may alter nutrient release across the GIT and influence enteroendocrine secretion. Our ^1^H NMR profiling and quantification analysis identified key metabolites modulated by food structure-including carbohydrates, amino acids, bile acids, and SCFAs.

To further understand how food structure shapes metabolic exposure along the GIT, we examined the spatial and temporal peak concentrations of key metabolites across the small intestine. Food structure had a marked effect on the timing and regional distribution of peak metabolite concentrations along the small intestine. In the duodenum, trigonelline, a chickpea-derived marker, reached peak levels earlier in clustered cell (CC) meals, followed by broken cell (BC) and then single cell (SC) structures, reflecting differences in digestion and transit. Similar temporal patterns were observed for stachyose and raffinose. In the ileum, amino acids and SCFAs were already present in the fasted state, suggesting they may represent background microbial fermentation products prior to meal arrival. Postprandially, amino acid concentrations increased from approximately 240 minutes onward, particularly following CC and BC meals, likely due to the delayed delivery of nutrient-rich digesta. Bile acids peaked later than other metabolites, with SC and BC meals showing more defined peak timing, whereas CC meals produced a highly variable bile acid profile, indicative of heterogeneous microbial processing. These observations highlight the importance of food matrices in shaping not only the composition but also the timing of nutrient exposure throughout the gut. Together, these patterns demonstrate that food structure dictates both the site and timing of nutrient sensing, with downstream effects on glycaemic response and hormone secretion.

BC released maltose/glucose rapidly in the mouth, stomach and duodenum, whereas SC and CC increased carbohydrate contents in the ileum, explaining different glycaemia responses reported between intact vs broken meals ^25^. CC and SC generally released nutrients more slowly/gradually than BC, which may explain the prolonged satiety of intact meals reported previously^1,26^. SC led to higher waves of SCFAs in the ileum than BC, likely due to its increased nutrient availability in the ileum which interacts with the local microbiome and enhances fermentation. We also observed higher levels of conjugated bile acids (BAs) in SC/CC than BC in the duodenum and ileum. One possible explanation is the interaction between plant cell wall components and bile acids, which may enhance bile acid retention ^27^. BAs could activate TGR5 and stimulate GLP-1 secretion, which would improve glucose metabolism ^28^. These findings provide mechanistic insights into how different food structures shape distinct luminal metabolite environments and affect metabolism, particularly glycaemia and appetite regulation. These observations suggest that gut hormone secretion, particularly GIP release, is closely associated with the rapid availability of carbohydrate in the stomach and duodenum in the BC, whereas the second peak of PYY and GLP-1 are related to the availability of amino acids and carbohydrate in the ileum.

### We observed high variability in the metabolite flow across individuals, especially in microbiome-related metabolites and in the ileum region

This variability underscores the complexity of individual responses to dietary intake and the value of personalized nutrition approaches. The ileum, despite hosting a diverse array of bacterial species, has been less studied than the large intestine. Previous work has described the small intestinal microbiome primarily through fasted-state sampling and genus-level 16S rRNA analyses, limiting insight into dynamic changes or strain-level resolution ^6^. Our study overcomes these limitations by capturing real-time, strain-resolved microbial profiles during digestion. While our fasted-state ileal communities reflected expected taxa such as *Streptococcus* and *Haemophilus*, we extended this static picture by showing that ingestion of a meal rapidly alters the microbial landscape, including the transient enrichment of oral-derived strains. These findings reveal the small intestine as a functionally dynamic environment, where microbial composition and activity shift in response to food structure and timing. In the fasted state, our results are comparable to other studies, with a community dominated by *Streptococcus* and *Haemophilus*, but containing a tightly associated community of species more associated with the colonic microbiome ^6,7^.

### Our findings significantly extend this view of the ileal microbiome to encompass dynamics during ingestion of a meal

Data from mouse models has suggested that the small intestinal microbiome is cyclical and responds to meals ^29^, but it was not clear if the same occurs in humans. We demonstrate that ingestion of a meal results in what in the literature was considered to be an “ileal segment specific microbiome” ^6^ being replaced by a microbiome which we have demonstrated through strain sharing analysis to originate in the oral cavity and to be enriched in strains identified in the saliva of the participants. We propose that these microbes are carried into the ileum via the bolus, where they are metabolically active. Previous studies have indicated that in ileostomy patients there may be links between a dynamic microbial community and metabolite profiles in the ileum ^30^, and in the ileum of healthy participants taking a probiotic supplement ^31^. Here we demonstrate that associations form between abundance of oral microbial strains and microbially derived metabolites, which represent the strongest microbe-metabolite associations following passage of the food bolus.

### Furthermore, we identify associations between the abundance of oral microbial strains in the ileum and the release of PYY hormone

This suggests a previously unknown mechanism whereby microbes from the oral cavity travel within the bolus to the ileum where they influence the molecular environment of the ileum and hormone release.

## Supporting information

Study protocol

Publication and Licensing Rights for Figure 1a

## METHODS

### Study scope, aims and structure/ Human study methodology

Our laboratory has made significant progress in addressing the gaps identified above, with *in vivo* sampling of the small intestine of healthy individuals by using nasoenteric tubes to retrieve the chyme from intact regions of the GI tract ^19,32^. Most recently, these samples have been analysed using ^1^H NMR, 16S sequencing and qPCR to study hormone release and microbial modifications of bile acids brought about by diet ^16^. Within this paper we have leveraged ^1^H NMR and shotgun sequencing to develop a pipeline characterising temporal profiles of metabolite bioavailability and high-resolution microbiome dynamics in nutrient-matched yet matrix-varied food, across the whole length of the intact small intestine.

This study aims to investigate the change in the molecular luminal environment over time and the role food structure plays in shaping this environment across the gastric, duodenal and ileal tract. To do so we tracked digestion of chickpeas in various forms (intact cell clusters, intact single cells, and broken cells), through the human gut using nasoenteral feeding tubes to gather synchronous samples of the gastric, duodenal, and ileal contents across 12 healthy overnight-fasted volunteers. ^1^H NMR of the samples were used to identify a variety of 35 metabolites commonly perturbed during digestion. These included amino acids, bile acids, short chain fatty acids (SCFAs), simple and complex carbohydrates. Given that the ileum has the highest bacterial concentration across the entire small intestine, shotgun sequencing of the ileal samples was also performed. This allowed for a microbiome-metabolome correlation analysis, which examined the relationship between food processivity and microbial populations.

### Test meals

Chickpea materials were provided by Quadram Institute Bioscience (Norwich Research Park, Norwich). Chickpeas were soaked and cooked using a Vorwerk Thermomix Version 5, following a procedure (Supplementary Text 1) to achieve three structures: broken cells, intact single cells, and cell clusters. The cooked chickpeas were mixed with 15 grams of Stutes no sugar-added blackcurrant jam and 115g of Harley’s no added-sugar raspberry jelly to add flavour. Each test meal provided 41.16 g available carbohydrate, 6.58g dietary fiber, 11.02g protein, 3.74g fat for a total 246 kcal. The amount of water served was adjusted to achieve the same portion size (760g).

### Human study design

The study was approved by the Health Research Authority and London-Camden and King’s Cross Research Ethics Committee (REC 19/LO/0962). The study has been prospectively registered at ISRCTN (https://www.isrctn.com/ISRCTN18097249, registration submitted on 22 Jannary 2020). Ten healthy participants aged 18–65□years with a BMI of 18.5–30□kg□m^2^ were recruited from the healthy volunteer database of NIHR Imperial Clinical Research Facility, based on the inclusion and exclusion criteria (Supplementary Table 1). Participant characteristics are provided in Supplementary Table 2. The protocol included one 4-day visit (phase 1), three 3-day visits (phase 2), and a 1-hour study visit (phase 3) at NIHR Imperial Clinical Research Facility (CRF) at Hammersmith Hospital. A minimum wash-out period of 7 days was required between each main visit to avoid cross-over effects. A consort diagram showing participant flow through the study is provided in Supplementary Figure 1.

The study followed a double-blinded, randomized crossover design. Randomization was performed by an independent researcher using a sealed envelope system (Sealed Envelope 2022). Both investigators and participants remained blinded until the completion of the study and data analysis.

As no prior studies had investigated the spatiotemporal metabolome and microbiome of the human gut in regard to food structure, sample size calculation was not feasible. Using similar intubation methods, Petropoulou et al.^19^ and Dagbasi et al.^16^ investigated duodenal and ileal metabolite profiles, respectively, observed significant differences between dietary interventions with 10 subjects. Thus, ten participants were recruited for this study.

### Phase 1 study – collection of gastric and duodenal samples

Ten participants attended a 4-day inpatient visit at NIHR Imperial Clinical Research Facility (CRF). The day before the study visit, participants were asked to refrain from caffeine, alcohol, and strenuous exercise. Participants were also requested to fast overnight. On day 1, participants received two enteral feeding tubes placed in their stomach and small intestine (duodenum) respectively. The placement of naso-enteric tubes for gastric and duodenal sampling was checked using the CORPAK (MedSystems, Halyard) feeding tube model that tracks the position of the tube during placement without the need for X-rays. On days 2,3, and 4, participants received three chickpea test meals in a randomized order. Gastric and duodenal samples were taken before and after the test meals for 180 min (T□=□−10, 0, 15, 30, 45, 60, 90, 120, 150 and 180□min). Blood samples were collected through a cannula placed in the antecubital fossa used for measuring gut hormone GIP, GLP-1 and PYY. On day 4, enteral tubes were removed, and participants were discharged. All participants adhered the protocol and received intended interventions.

### Phase 2 study-collection of ileal samples

Two participants withdrew the phase 1 study, (one due to the burden of intubation and one due to the time commitment), leaving eight participants who entered the phase 2 trial. Eight participants attended three 3-day inpatient visits at NIHR Imperial CRF. Participants were asked to refrain from caffeine, alcohol, and strenuous exercise before attending the visits. Participants were also requested to fast overnight. On each visit, participants received one of these chickpea-based meals, in randomised order. On day 1, a nasoenteric tube was inserted into the participants’ terminal ileum. The tube position was validated using fluoroscopy ^32^ by professional radiologists at Charing Cross Hospital. Day 2 allowed participants to acclimatize to the diet and environment. On day 3, ileal samples were collected before breakfast and every 60 min until 480 min. Blood samples were collected at the same intervals for measuring gut hormone GLP-1 and PYY. All eight participants adhered the protocol and received intended interventions.

### Phase 3 study-Collection of oral samples

The recruitment target for the phase 2 study was 10 participants. Those who completed phases 1 and 2 were preferentially invited to take part in phase 3. Eight participants responded and attended a one-hour visit at the NIHR Imperial CRF. In addition, two new participants were recruited from the healthy volunteer database of NIHR Imperial CRF to take part in the phase 3 visit only. During the visit, participants were instructed not to brush their teeth or use mouthwash for at least two hours before arriving. They were asked not to consume any food, drink, chewing gum or tobacco. 2 ml of saliva was collected as a baseline.

The chickpea meals were prepared using the same protocol as in phase 1 and 2 and then weighed out to approximately 12-18 g to achieve equivalent macronutrient content (each meal provided 1.0 g of starch). Participants were asked to chew the chickpea meals, served in a randomised order, to the urge to swallow, stop chewing and provide us with oral samples. The oral samples were immediately stored at −80 degrees for further analysis. All ten participants adhered the protocol and received intended interventions.

### Metabolite extraction

Oral, gastric, duodenal and ileal samples were centrifuged at room temperature for 15 minutes at 3000g to remove particulates. The resulting supernatant was collected and 450ul of the supernatant was used for metabolite extraction. Metabolites were extracted by adding 1ml methanol, 2ml chloroform, and 1 ml water (ratio 1:2:1). Samples were vortexed for 1 minute and centrifuged at 4 degree for 20 minutes at 3000g. After centrifugation, samples were separated into aqueous and the organic phase. The aqueous phase was evaporated in a Speed Vacuum concentration (Eppendorf Concentrator Plus) to dryness and stored at –80 °C for further analysis.

### NMR spectroscopic analysis

The aqueous phase of gut samples was re-constituted in 640 μL of H2O and sonicated for 10 minutes. Samples were checked after sonication and there was no visible precipitation. 60 μL of NMR buffer [1.5 M KH2PO4 buffer (pH 7.4, 100% of deuterium oxide (D_2_O), 2 mM sodium azide and 1 mM TSP (3-trimethylsilyl-[2,2,3,3,-2H4]-propionic acid sodium salt)] was mixed with 540 μL of samples and the mixture was further transferred to 5 mm NMR tubes. Quality controls were prepared by pooling 100 μl of each sample, independently for gastric and duodenal samples.

^1^H NMR spectroscopy was performed at 300 K on a Bruker 600 MHz spectrometer 66 (Bruker Biospin, Karsruhe, Germany). It followed a standard one-dimensional pulse sequence with saturation of the water resonance RD – gz,1 – 90° – t – 90° – tm – gz,2 – 90° – ACQ (noesygppr1d) where RD is the relaxation delay, 90° represents the applied 90° radio frequency (rf) pulse, t is an interpulse delay set to a fixed interval of 4 μs, RD was 2 s and tm (mixing time) was 100 ms. Water suppression was achieved through irradiation on the water signal during RD and tm. Each 21 spectrum was acquired using 4 dummy scans followed by 64 scans and collected into 64 K data points. The receiver gain was 90.5. A spectral width of 20,000 Hz was used for all the samples. Prior to Fourier transformation, the FIDs were multiplied by an exponential function corresponding to a line broadening of 0.3 Hz.

### NMR data processing

Multivariate statistical analysis was performed on spectral data. Each spectrum (∼24 K spectral variables) was automatically phased, baseline corrected, digitized over δ −0.5 to 10 and imported into MATLAB (R2021a, Mathworks, Inc.; Natwick, USA), using in-house scripts. Spectral regions corresponding to the internal standard (δ −0.5 to 0.5) and water (δ 4.7 to 4.9) were excluded.

### Metabolite identification

Statistical spectroscopic tools such as Statistical Total Correlation Spectroscopy (STOCSY) and Subset optimization by reference matching (STORM) were used to identify spectra signals. Internal and external databases such as the Human Metabolome Data Base (HMDB, Human Metabolome Data Base (HMDB; http://hmdb.ca/) and the Biological Magnetic Resonance Data Bank (BMRB; http://www.bmrb.wisc.edu) were used for assignment purposes. Arguably, these assignments have not been confirmed as it would require spiking.

### Metabolite quantification

For metabolic profiling, we chose 35 metabolites to quantify, based on the most likely metabolites encountered in consumed legumes. These include 5 sugars (stachyose, raffinose, maltose, glucose), metabolic acids like SCFAs (n-butyrate, propionate, formate, fumarate, acetate), 9 amino acids (valine, alanine, glutamine, methionine, histidine, aspartate, asparagine, tyrosine, phenylalanine), 6 bile acids (TCA, GCA, TDCA, GDCA, TCDCA, GCDCA) and metabolic markers of chickpeas such (trigonelline, tryptophan, ciceritol).

During meal preparation, metabolites may be affected by processing loss (mechanical damage, leaching), thermal degradation, enzymatic degradation, oxidation, prolonged storage times and complex chemical interactions. Hence, we have built in multiple layers of redundancy, by tracking molecule families. For example, in addition to using trigonelline, tryptophan and ciceritol as metabolic markers for chickpeas, we also used stachyose and raffinose to track the passage of chickpeas through the GI-tract. These two small soluble cell wall components are resistant to small digressional enzymes but are released rapidly in the oral cavity. Furthermore, for the GI-samples it was easier to robustly quantify stachyose-raffinose as a joint peak in the NMR data as opposed to two separate molecules.

The quantification of metabolites was achieved using in-house developed routines written in both JavaScript and R. NMR spectra were processed according to the Bruker IVDr standard procedures ^33^. No further processing deemed necessary. Normalisation is not required prior to quantification and can be applied on the extracted concentrations when necessary. The following steps were followed for quantification:

1. The metabolites of interest were grouped in spectral regions of interest (SROI).
2. For each SROI, a list of signals was established with attributes that define their characteristic patterns (chemical shift, linewidth, multiplicity and scalar coupling). All the patterns present in an SROI were included, even for unknown signals.
3. Those parameters were used as initial input for a gradient descent optimization (Levenberg-Marquardt) algorithm to minimize the difference between the experimental and the fitted pattern.
4. The resulting models were graded using a comprehensive heuristic, including metrics specific to the optimisation error and metrics capturing the correlation of integrals registered for patterns known to belong to the same compound.
5. Models with lower grades were visually inspected using a suite of interactive web tools build in JavaScript ^34^.
6. The resulting areas under the curve were then converted to concentrations using the ERETIC internal standard and calibration procedure ^35^ that is part of the Bruker IVDr protocol.

Steps 1-2 were repeated for the 2 different sample matrix type, i.e., for both the ileum and chickpea samples.

### Metabolite Modelling

Quantification of metabolites from the proton nuclear magnetic resonance (^1^H NMR) spectroscopy was performed^1^ and metagenomic sequencing of the microbiome were performed. However, metagenomic sequencing was restricted to ileal samples, as gastric samples were expected to be sterile, and duodenal samples, though processed, exhibited extremely low microbial abundance and were consequently discarded.

For metabolite analysis, data imputation was performed using the MissForest algorithm to account for inconsistent timepoints across the 10 gastric, 10 duodenal, and 8 ileal participants. Oral samples were excluded from this imputation due to extreme data sparsity, with only 8 oral metabolites quantified compared to 28 in the gastric, duodenal, and ileal dataset. Additionally, oral metabolite profiles differed substantially from gastric timepoint 0, and data were available for only 8 of the 10 volunteers. Nevertheless, oral samples were analyzed separately, and two additional participants were included to compensate for missing data points in this subset.

Principal component analysis (PCA) (Fig. 1b) was conducted following imputation, confirming the absence of participant outliers. Metabolite concentrations were averaged across all participants per timepoint and diet, and the resulting means were visualized in three-dimensional graphs (Fig. 2). Box-and-whisker plots, available in the Supplementary Information, provide further details on participant variance. Pairwise comparisons between meals were performed using dynamic time warping (DTW) (Fig. 9), with the Broken Cells (BC) diet serving as the reference. A smaller DTW distance indicates a higher similarity between the BC diet and the diet being evaluated (SC or CC).

The GI location (gastric, duodenal, ileal) and timepoint of peak concentration for each metabolite, averaged across metabolite families within each diet, were identified and represented in a scatter plot (Fig. 3). This qualitative visualization highlights the temporal and spatial distribution of peak concentrations for amino acids, bile acids, chickpea markers, fatty acids, and sugars following ingestion. Finally, oral sample data (Fig. 2, box-and-whisker plots) were analyzed separately from the gastric, duodenal, and ileal dataset to illustrate inter-individual variability, upper and lower concentration limits, and median concentrations.

### DNA extraction and metagenomic sequencing

DNA extraction and metagenomic sequencing was only done for ileal samples, as gastric samples were too acidic and duodenal sample quality was too low to retrieve metagenomes. DNA extraction was carried out per the manufacturer’s instructions with the MP Bio fast DNA spin kit for soil (MP Biomedical, Solon, USA). Ileal aspirates were transferred to Lysing Matrix E tubes where 980 µL of sodium phosphate buffer and 120□µL of MT buffer were added. The samples were homogenised in the FastPrep24 bead-beating instrument (MP Biomedicals, Solon USA) for 3□min (3 runs of 60□s each, with 5□min rest on ice in between). Afterwards, samples were centrifuged at 14,000□×□*g* for 15□min, and the sample supernatant was transferred into clean 2□mL microcentrifuge tubes with 250□µL of protein precipitate solution. The tubes were mixed by shaking by hand 10 times before centrifugation at 14,000□×□*g* for a further 10□min. The supernatant was transferred to 5□mL tubes with a resuspended binding matrix (1□mL of silica slurry that binds DNA from lysates), and the tubes were inverted for 2□min allowing the DNA to bind. The tubes were placed in a rack and allowed to settle for 1□h. Then, 500□µL of the supernatant was removed, and the binding matrix was resuspended in the remaining supernatant. The washing steps involved the transfer of 700□µL of the mixture to a SPIN filter centrifuged at 1000□×□*g* for 2□min to empty the catch tube and transfer the leftover mix. This process was repeated until all the mixture had been added to the SPIN filter. The SPIN filter was washed with 500□µL of ethanol-based wash solution SEWS-M. The samples were centrifuged at 14,000□×□*g* for 5□min, and the catch tube was emptied. Then, samples were centrifuged a second time (Dry spin) at 14,000□×□*g* for 5□min to remove the residual wash solution. The catch tube was replaced with a clean tube (1.5□mL LoBind Eppendorf^®^ tubes), and samples in the SPIN filter were air-dried for 10□min at room temperature and 5□min at 37□°C. Then, 65□µL of DNA extraction solution was added to the Binding Matrix and incubated at room temperature for 5□min before being centrifuged at 6600□×□*g* for 2□min with lids open to bring eluted DNA into the tube. The SPIN Filter was discarded, and the eluted DNA was stored at −20□°C.

Genomic DNA was normalised to 5□ng/µL with elution buffer (10□mM Tris HCl). A miniaturised reaction was set up using the Nextera DNA Flex Library Prep Kit (Illumina, Cambridge, UK). 0.5□µL Tagmentation Buffer 1 (TB1) was mixed with 0.5□µL Bead-Linked Transposomes (BLT) and 4.0□µL PCR-grade water in a master mix and 5□µL was added to each well of a chilled 96-well plate. About 2□µL of normalised DNA (10□ng total) was pipette-mixed with each well of Tagmentation master mix and the plate heated to 55°C for 15□min in a PCR block. A PCR master mix was made up using 4□µL kapa2G buffer, 0.4□µL dNTP’s, 0.08□µL Polymerase and 4.52□µL PCR-grade water, from the Kap2G Robust PCR kit (Sigma-Aldrich, Gillingham, UK) and 9□µL added to each well in a 96-well plate. About 2□µL each of P7 and P5 of Nextera XT Index Kit v2 index primers (catalogue No. FC-131-2001 to 2004; Illumina, Cambridge, UK) were also added to each well. Finally, the 7□µL of Tagmentation mix was added and mixed. The PCR was run at 72□°C for 3□min, 95□°C for 1□min, 14 cycles of 95□°C for 10□s, 55□°C for 20□s and 72□°C for 3□min. Following the PCR reaction, the libraries from each sample were quantified using the methods described earlier and the high sensitivity Quant-iT dsDNA Assay Kit. Libraries were pooled following quantification in equal quantities. The final pool was double-SPRI size selected between 0.5 and 0.7X bead volumes using KAPA Pure Beads (Roche, Wilmington, US). The final pool was quantified on a Qubit 3.0 instrument and run on a D5000 ScreenTape (Agilent, Waldbronn, DE) using the Agilent Tapestation 4200 to calculate the final library pool molarity. qPCR was done on an Applied Biosystems StepOne Plus machine. Samples quantified were diluted 1 in 10,000. A PCR master mix was prepared using 10□µL KAPA SYBR FAST qPCR Master Mix (2X) (Sigma-Aldrich, Gillingham, UK), 0.4□µL ROX High, 0.4□µL 10□μM forward primer, 0.4□µL 10□μM reverse primer, 4□µL template DNA, 4.8□µL PCR-grade water. The PCR programme was: 95□°C for 3□min, 40 cycles of 95□°C for 10□s, 60□°C for 30□s. Standards were made from a 10□nM stock of Phix, diluted in PCR-grade water. The standard range was 20, 2, 0.2, 0.02, 0.002, 0.0002□pmol. For the ileal samples the pooled library was then sent to Novogene (Cambridge, UK) for sequencing using an Illumina NovaSeq 6000 instrument, with sample names and index combinations used. Demultiplexed FASTQ’s were returned on a hard drive. A sequencing depth of ∼12.6GB per sample was achieved. For the oral samples, the pooled library was sequenced by Novogene (Cambridge, UK) using an Illumina Novaseq X instrument to a sequencing depth of ∼25GB per sample.

### Metagenomic data processing

The raw data were processed using the MG-TK pipeline (formerly MATAFILER) ^36^ Specifically, reads were filtered based on quality using sdm (simple demultiplexer). Reads matching the human or chickpea genome were identified with kraken2 and filtered out. Samples from the same individual were co-assembled using MEGAHIT and binned using SemiBin. Reads were mapped to the assembly with Bowtie 2. Genes were predicted with Prodigal and SNP calling was performed using bcftools. MAGs were clustered into metagenomic species (MGS) and then taxonomically profiled based on marker genes using GTDB-tk and LCA algorithm. For each high quality MGS a phylogenetic tree was calculated based on SNP-resolved gene orthologues using IQ-Tree. Phylogenetic trees were then used for strain-level analysis following procedure published previously ^17^. Briefly, this approach assumed a presence of one dominant strain per sample and a case of strain sharing was defined if a pair of samples from the same individual formed a monophyletic clade and the distances between them fell within tree-specific phylogenetic distance threshold. For each MGS and participant at a given timepoint, strain sharing ratio have been calculated as a proportion of samples with the same strain among all oral and ileal samples. The pipeline has been used an assembly dependent manner, so the counts are reads mapping to marker genes in metagenome assembled genome bins. Raw reads have been normalised for gene length/copy number.

Networks combining species and metabolite data for each time point were produced using SPIEC-EASI. Each network used parameters nlamba 100, lamba.min.ratio 1e-1, method “glasso”, sel.criterion “bstars”, and pulsar parameter rep.num 100. Species or metabolites which were observed in 20% or fewer of samples were filtered before network construction. For visualisation vertices with degree 0 were removed.

### Gut hormone assays

Plasma GIP concentrations were measured by Human GIP ELISA kits from (EZHGIP-54K, Merck). All these assays were performed as per the manufacturers’ instructions. GLP-1 and PYY concentrations were measured by the in-house RIA method^37^. Statistical differences between groups were analyzed using repeated measure two-way ANOVA followed by Tukey’s post-hoc test.

## ACKNOWLEDGEMENTS

The authors thank the research participants for their participation and providing samples in this trial. We thank Dr Martina Tashkova and Dr Shilpa Tejpa (Imperial College London) for their assistance with sample collection. The research was carried out at the NIHR Imperial Clinical Research Facility. This research was funded by the Biotechnology and Biological Sciences Research Council, UK, Institute Strategic Programme grant BB/R012512/1 and its constituent project BBS/E/F/000PR10345 and BB/X011054/1 and its constituent project BBS/E/F/000PR13630, and its partner project BB/X018857/1. M.C. is funded by the China Scholarship Council from the Ministry of Education of PR China for her PhD. E.H. holds an Australian Research Council Laureate Fellowship. The Western Australian State Government and the Medical Research Future Fund provided funding for the Australian National Phenome Centre. The funders had no role in study design, data collection and analysis, decision to publish or preparation of the manuscript.

## AUTHOR CONTRIBUTIONS

Conceptualization: G.F.; Methodology: F.J.W., D.B., I.G.P., C.E., F.H., E.H., J.W. and G.F.; Investigation: N.F., M.C., F.J.W., A.D., H.H., J.I.S.C., K.S., A.B., A.C., K.P., D.B. and N.P.M.; Formal analysis: N.F., M.C., F.J.W., A.D., H.H., J.I.S.C., K.S., A.B., A.C., N.P.M., I.G.P., C.E., F.H., E.H., J.W. and G.F.; Visualization: N.F., F.J.W. and G.F.; Supervision: F.J.W., I.G.P., C.E., F.H., E.H., J.W. and G.F.; Funding acquisition: G.F.; Writing – original draft: N.F., M.C., F.J.W. and G.F.; Writing – review & editing: all authors. N.F., M.C. and F.J.W. contributed equally to this work; I.G.P., C.E., F.H., E.H., J.W. and G.F. contributed equally to the generation of novel tools, analysis and construction of the manuscript.

## ADDITIONAL INFORMATION

Correspondence and requests for materials should be addressed to either Nadia Fernandes or Mingzhu Cai.

## DATA AVAILABILITY

The data reported in this study are available from the Mendeley Data Database at https://data.mendeley.com/preview/dnz2xpkgx3?a=8340a9d8-5a33-4eda-a4ff-89bd1edb95a7. Metagenomic sequencing data is available through the NCBI SRA through project code PRJNA1240928.

## CODE AVAILABILITY

Custom codes used in this study are available from https://github.com/Nerdobyte/Diet_Metabolome_Microbiome.

## EXTENDED FIGURES

**Extended figure 1:**
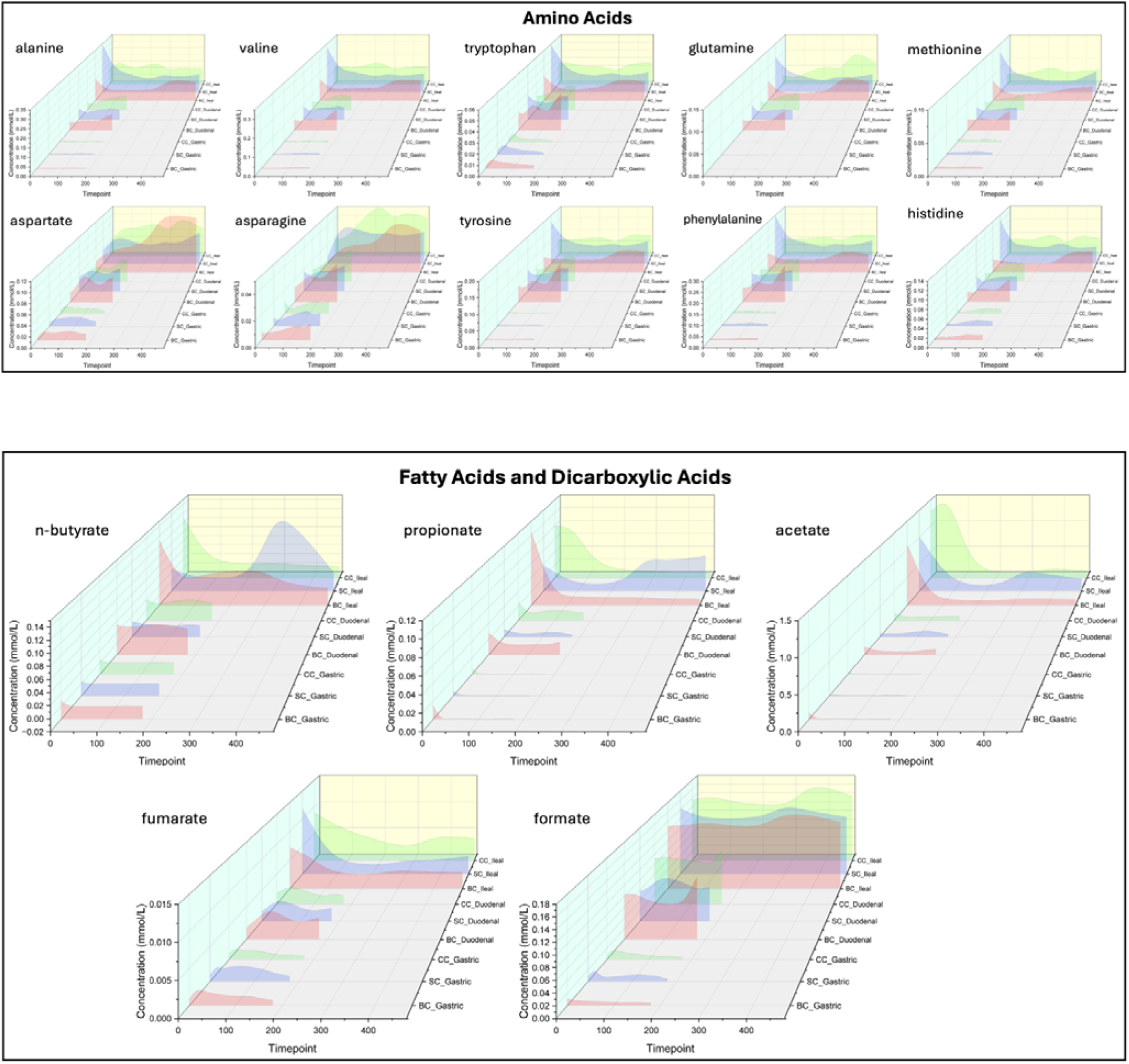
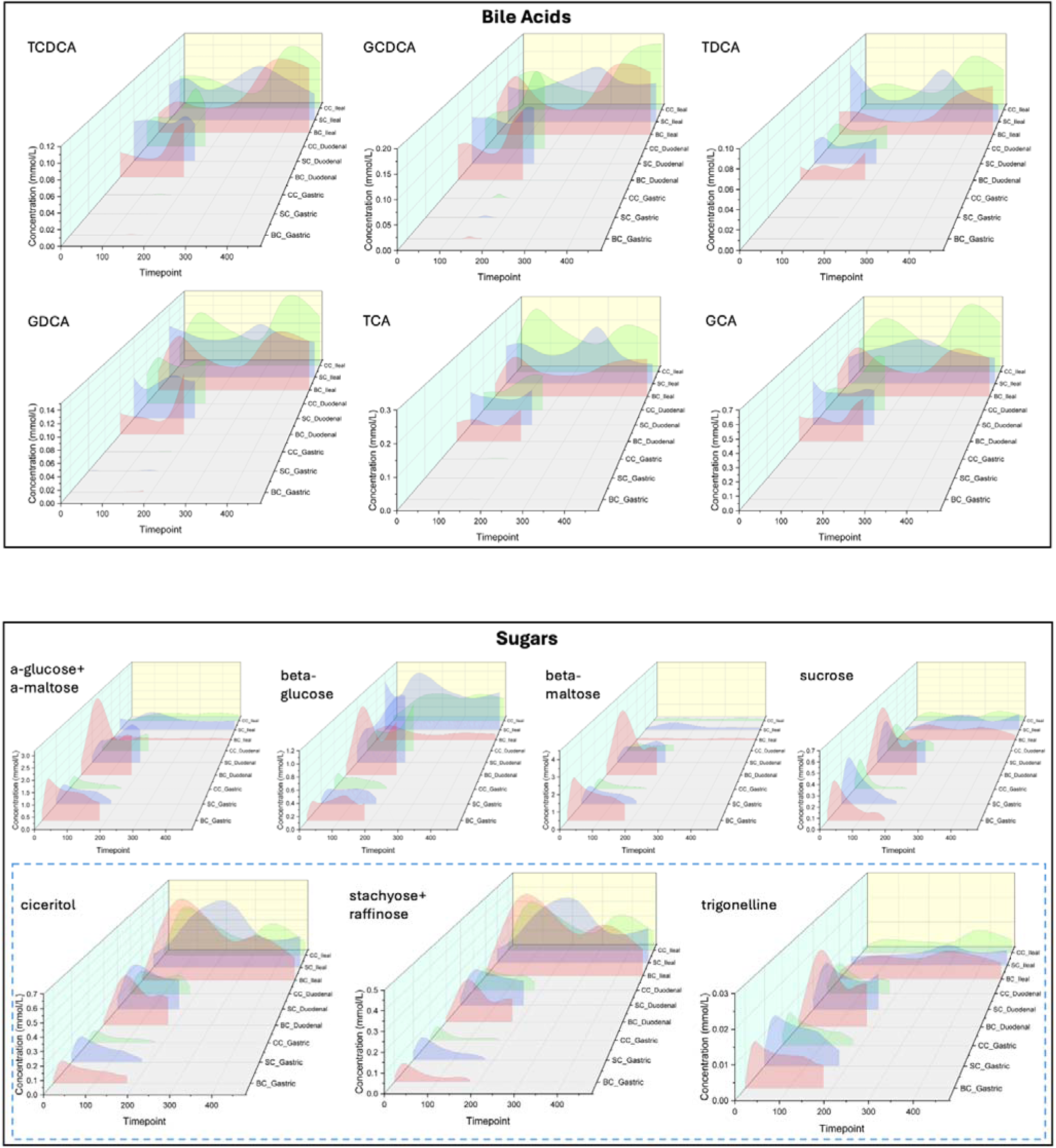
3D graph tracking of metabolite concentrations across 3 different diets along the GI-tract. Diet BC (pink) corresponds to broken cells, diet SC (blue) corresponds to single cells and diet CC (green) corresponds to cell clusters. Z-axis shows samples in the following order: three gastric samples, three duodenal samples and ileal samples. Inset blue box (dashed) within sugars are chickpea markers.

**Extended Figure 2:**
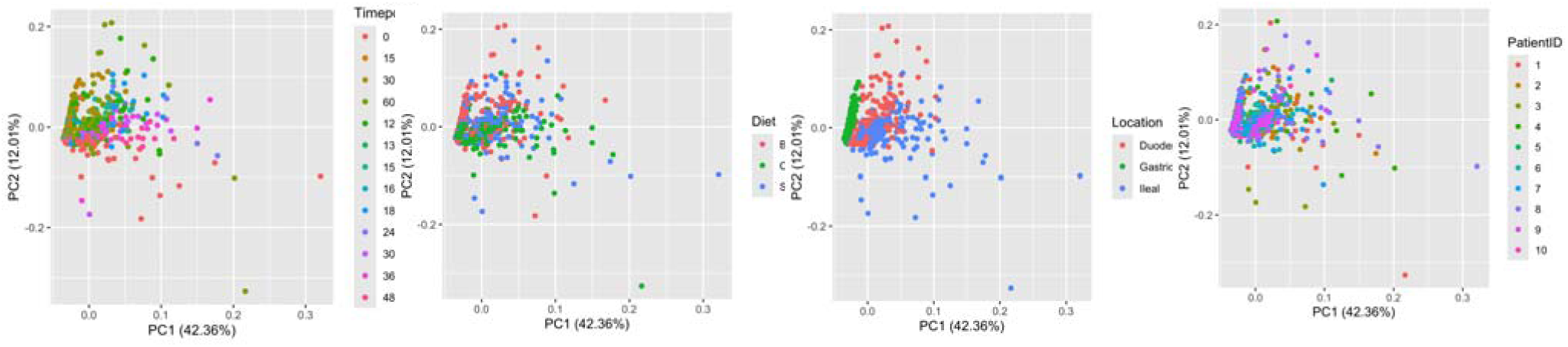
Principal Component Analysis of all metabolites at all timepoint, colored by timepoint, diet, location of sampling within the gastrointestinal tract and patient ID.

**Extended Figure 3:**
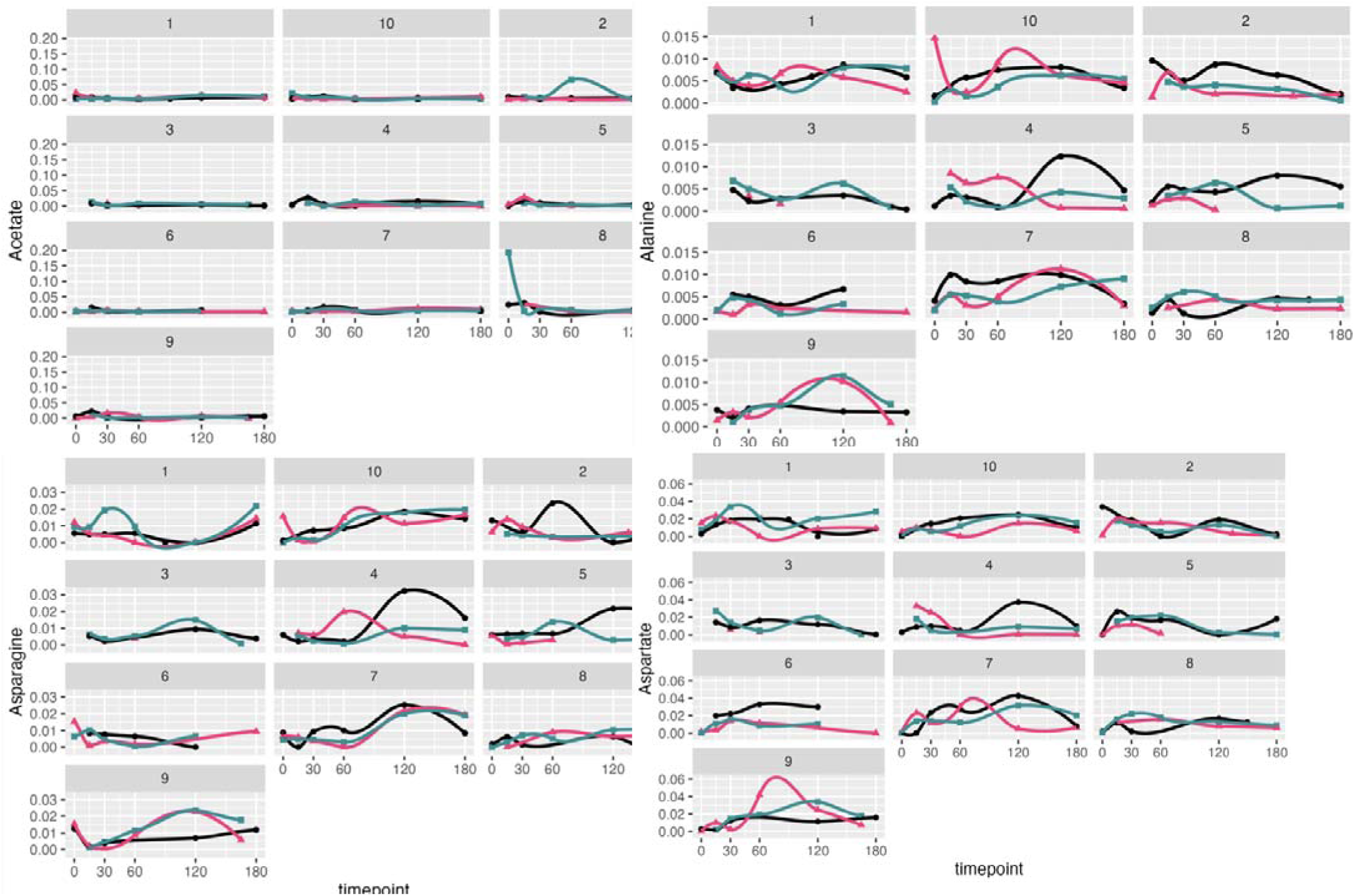
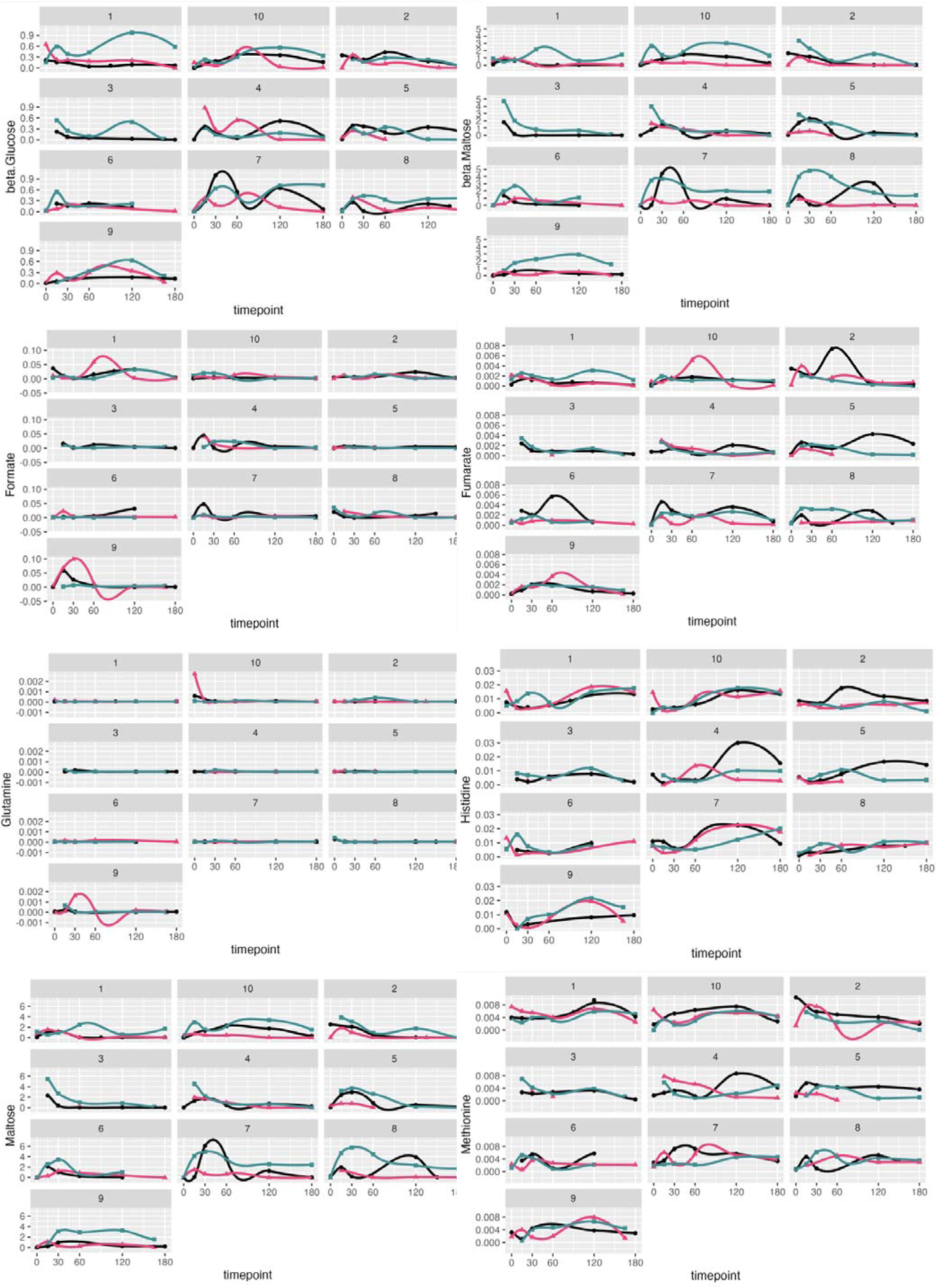
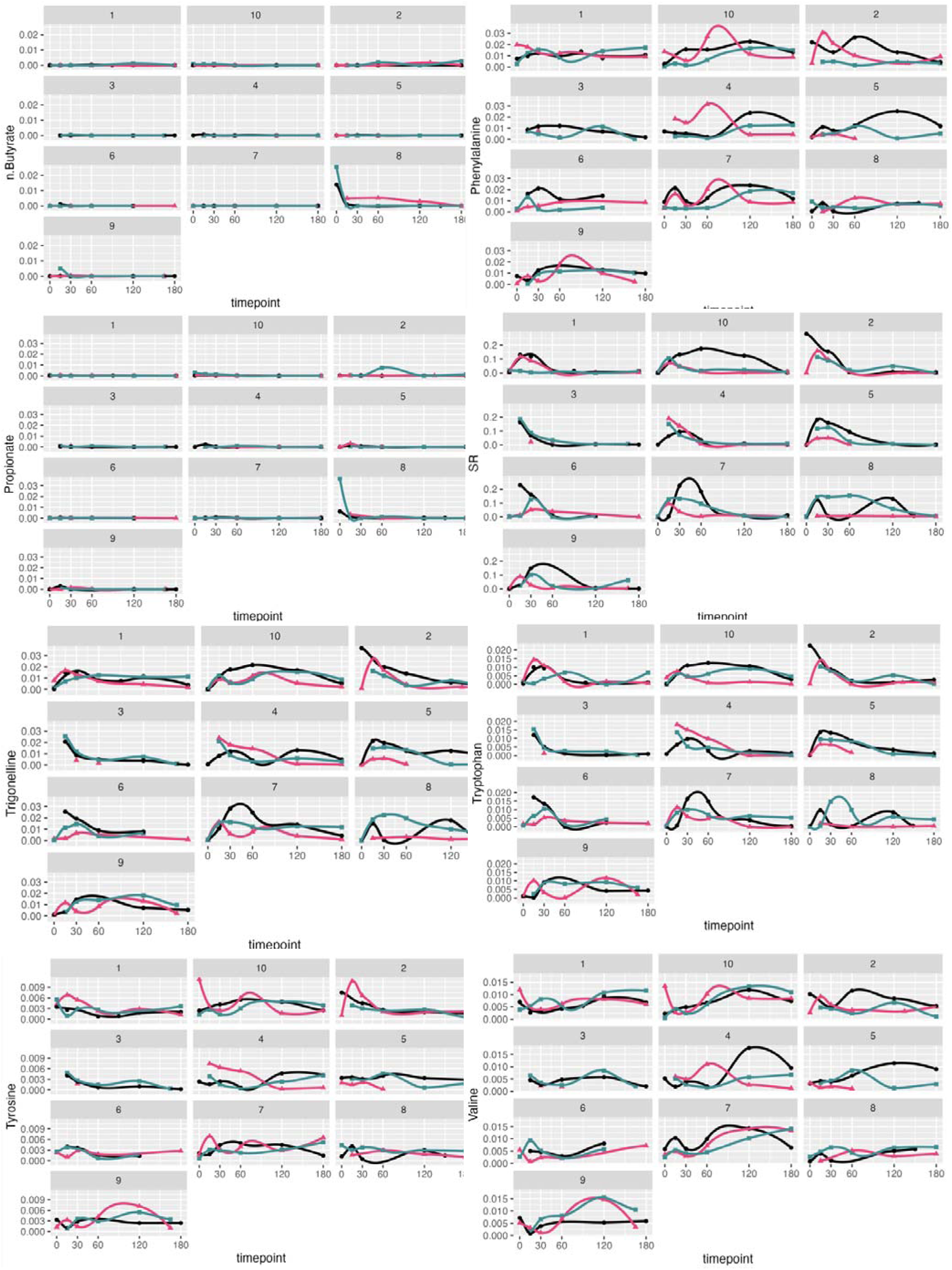
Per individual metabolic concentration in mmol/L plotted against time after meal consumption, within **gastric** samples. Green curves are diet BC, Red curves are diet CC, Black curves are diet SC.

**Extended Figure 4:**
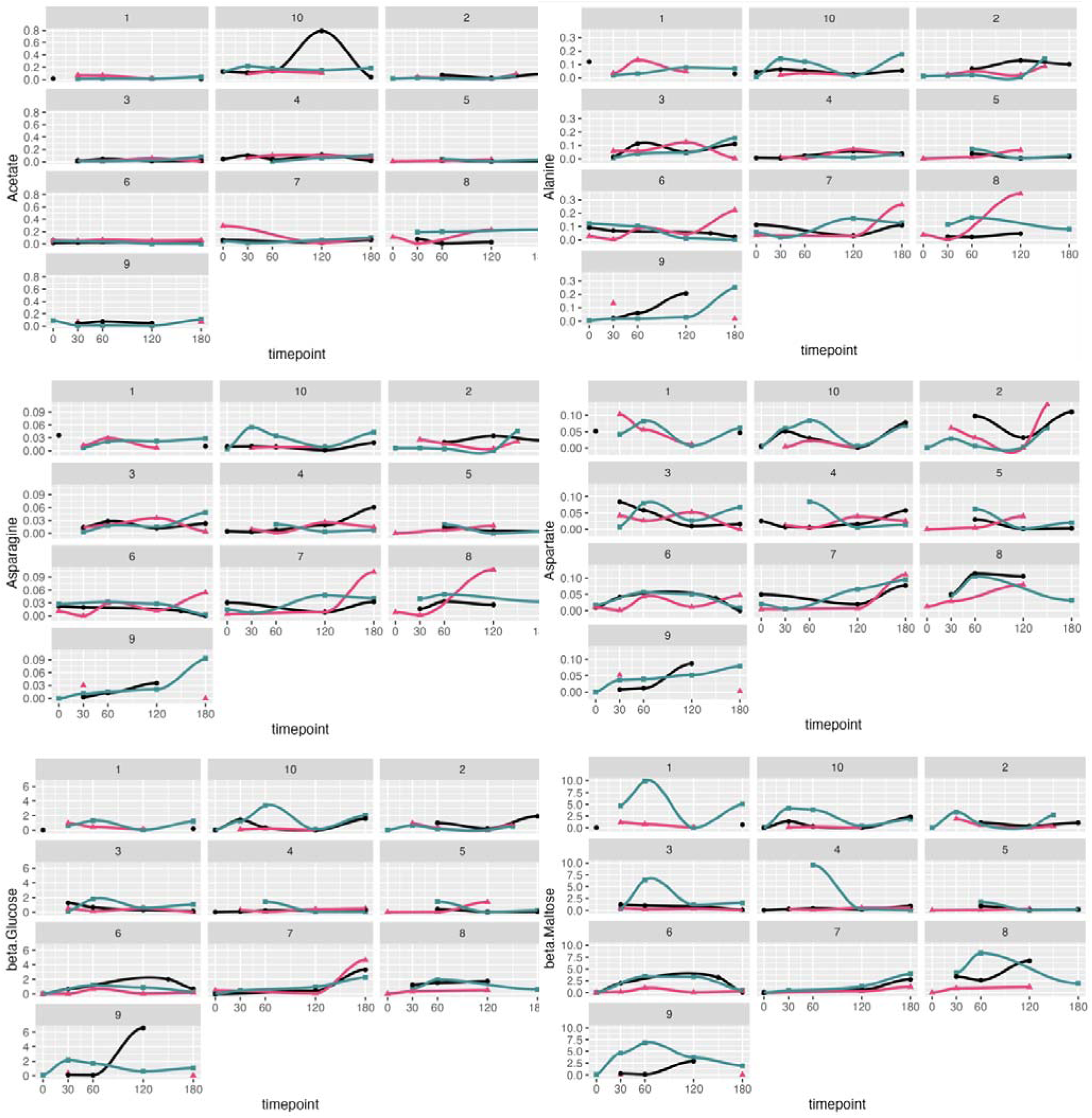
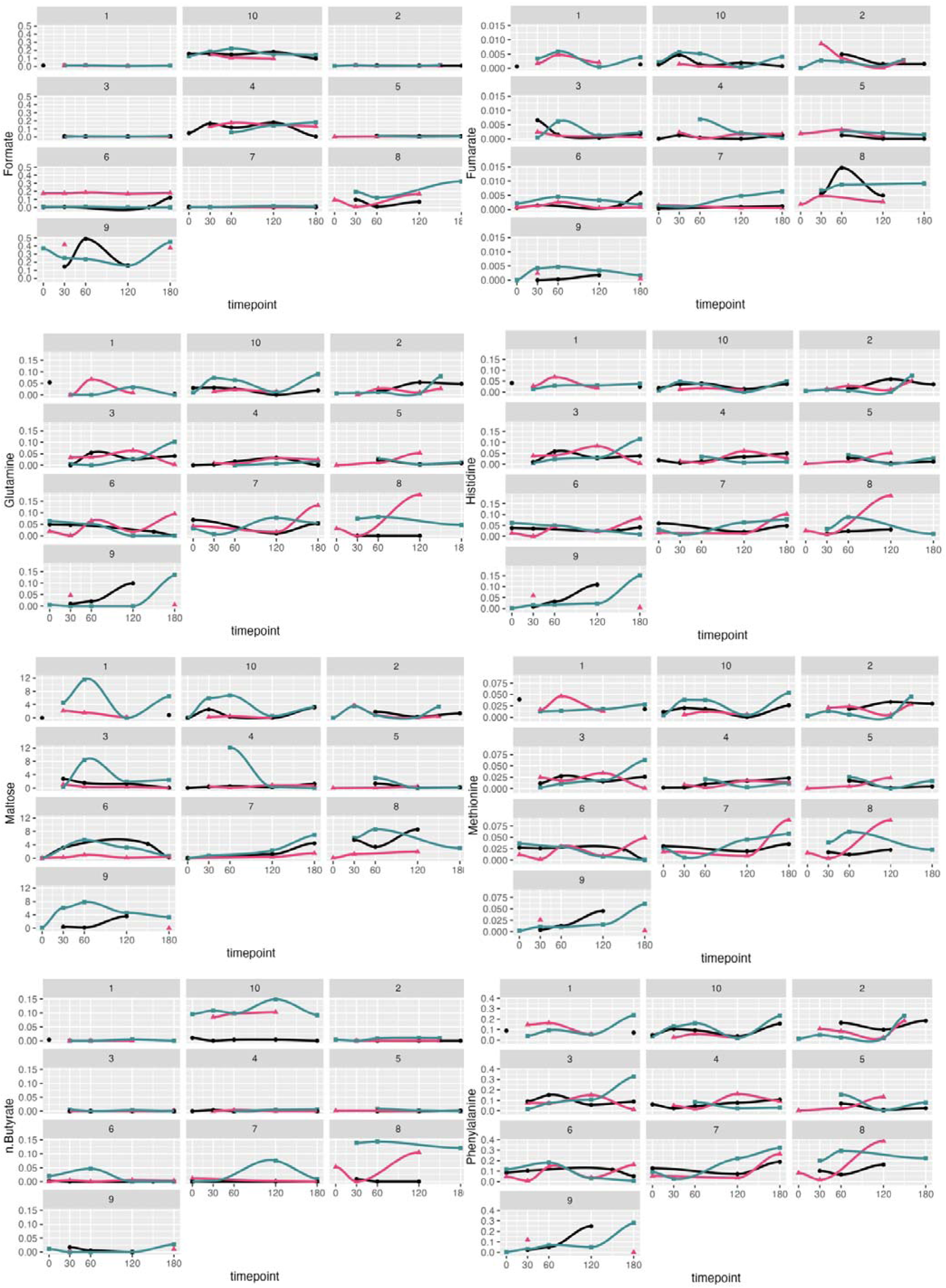
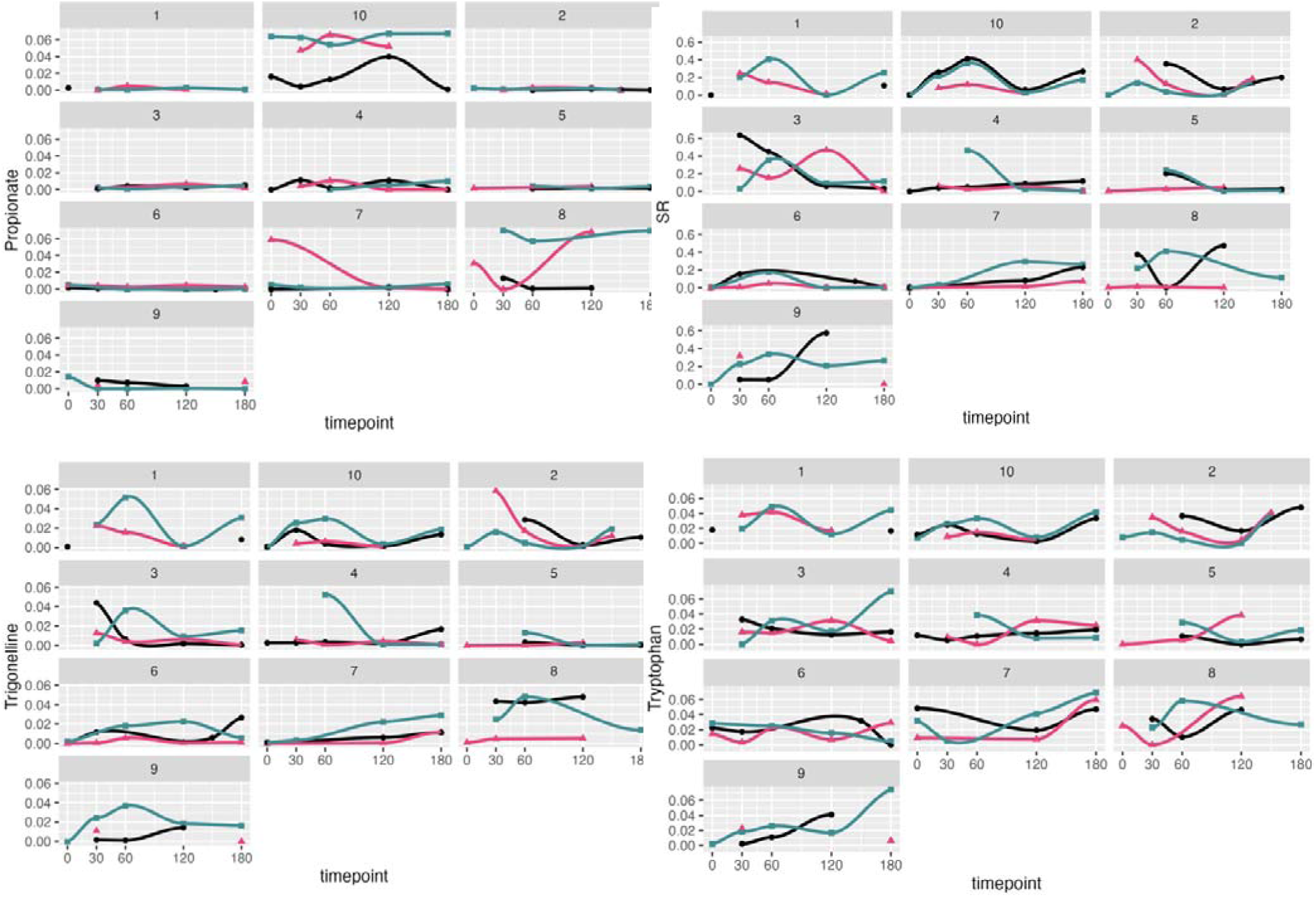
Per individual metabolic concentration in mmol/L plotted against time after meal consumption, within **duodenal** samples. Green curves are diet BC, Red curves are diet CC, Black curves are diet SC.

**Extended Figure 5:**
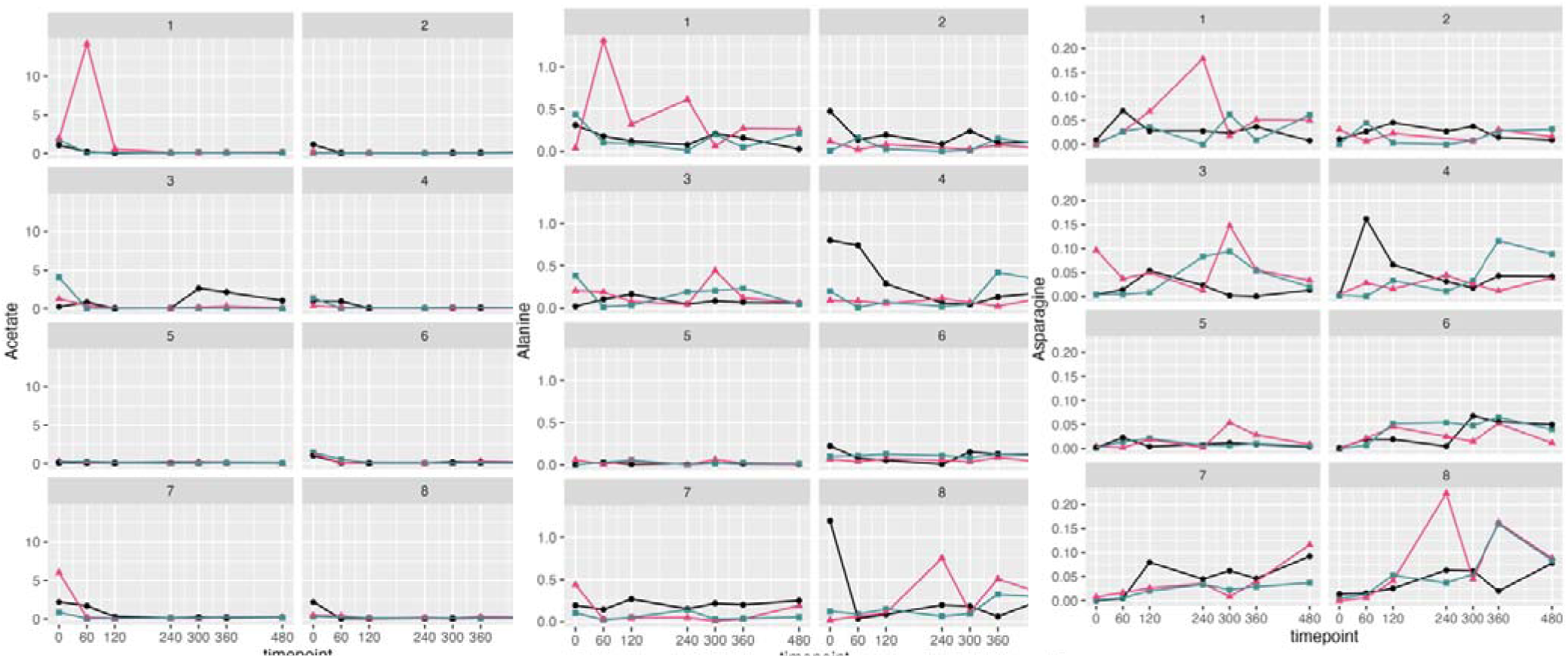
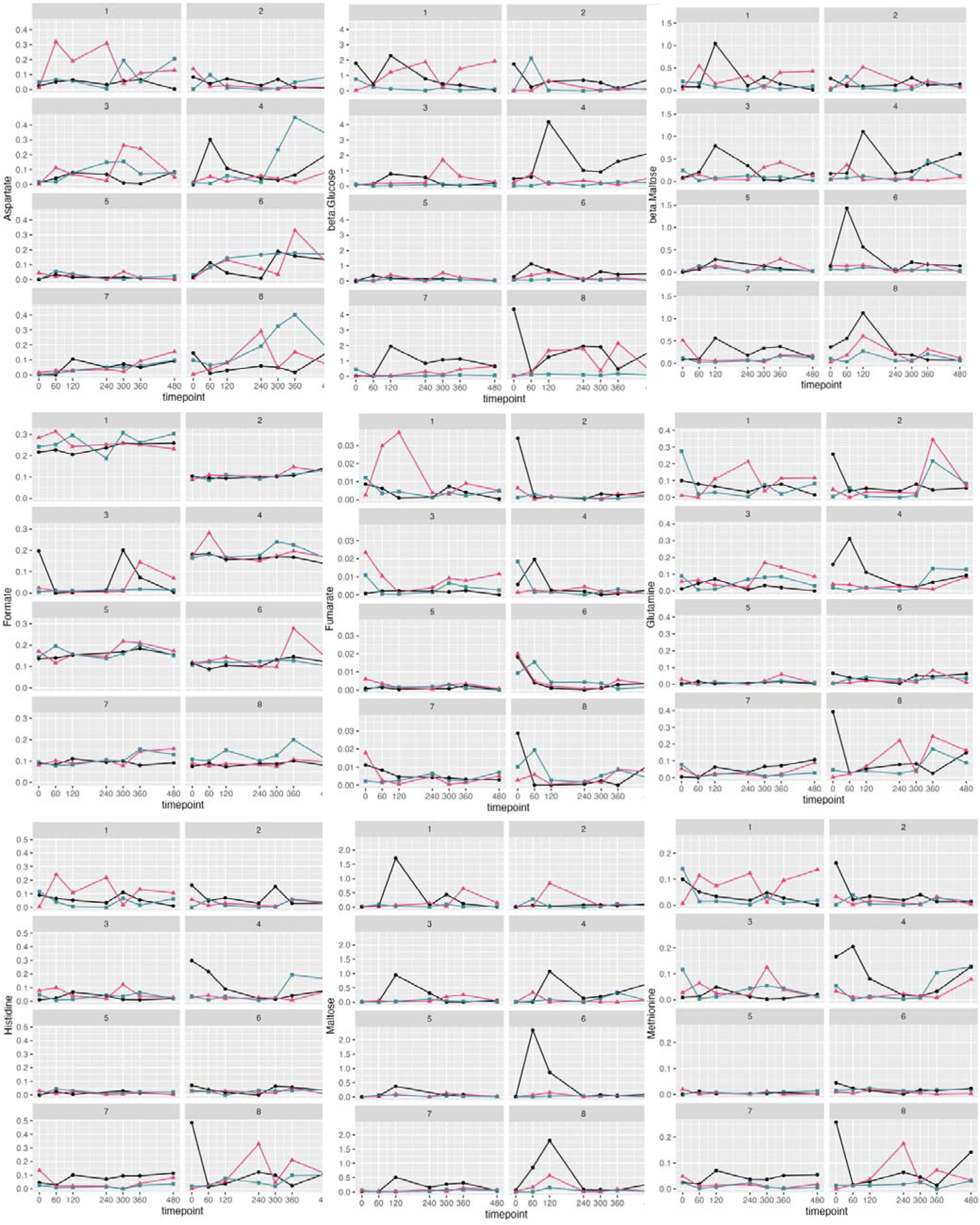
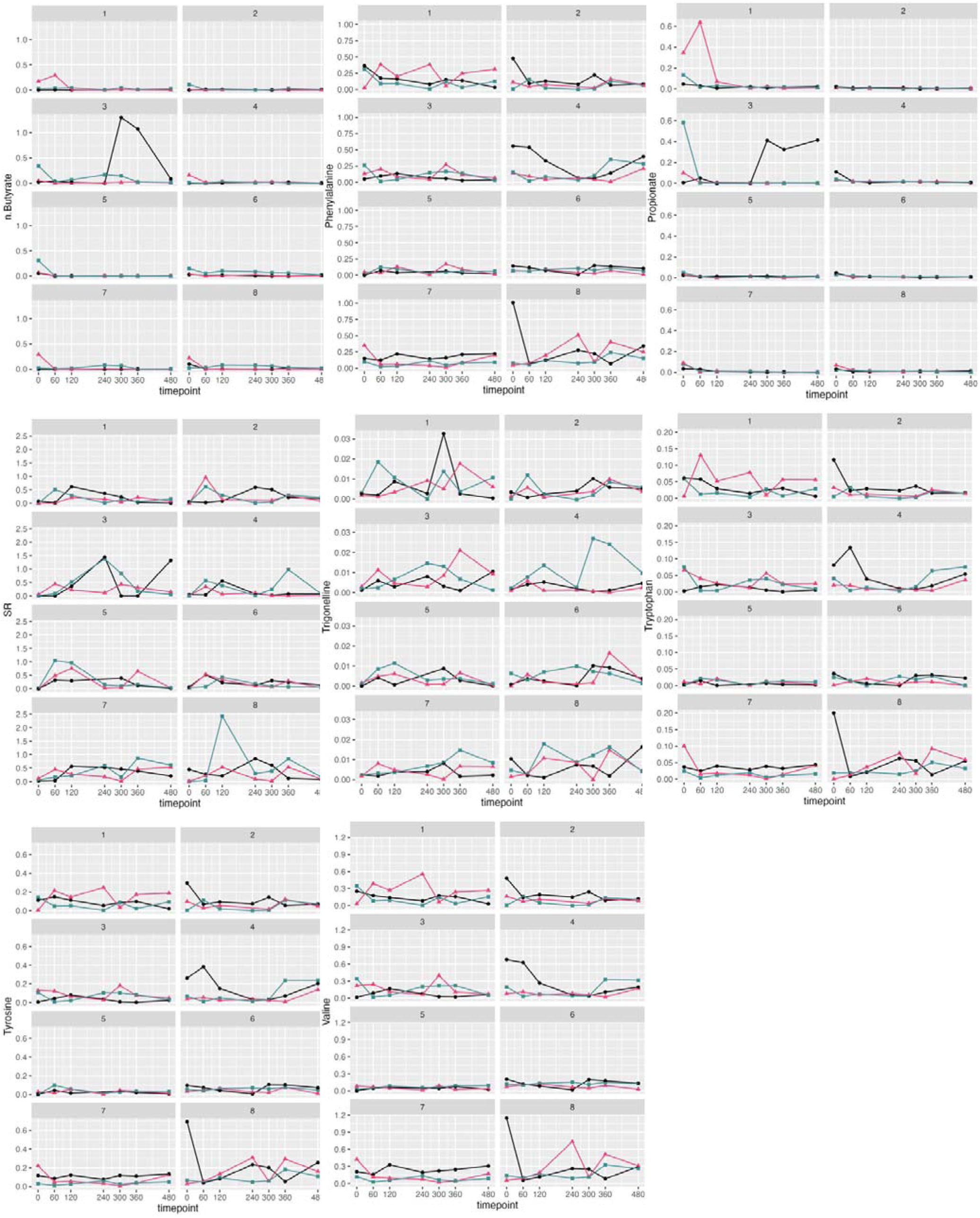
Per individual metabolic concentration in mmol/L plotted against time after meal consumption, within **ileal** samples. Green curves are diet BC, Red curves are diet CC, Black curves are diet SC.

**Extended Figure 6:**
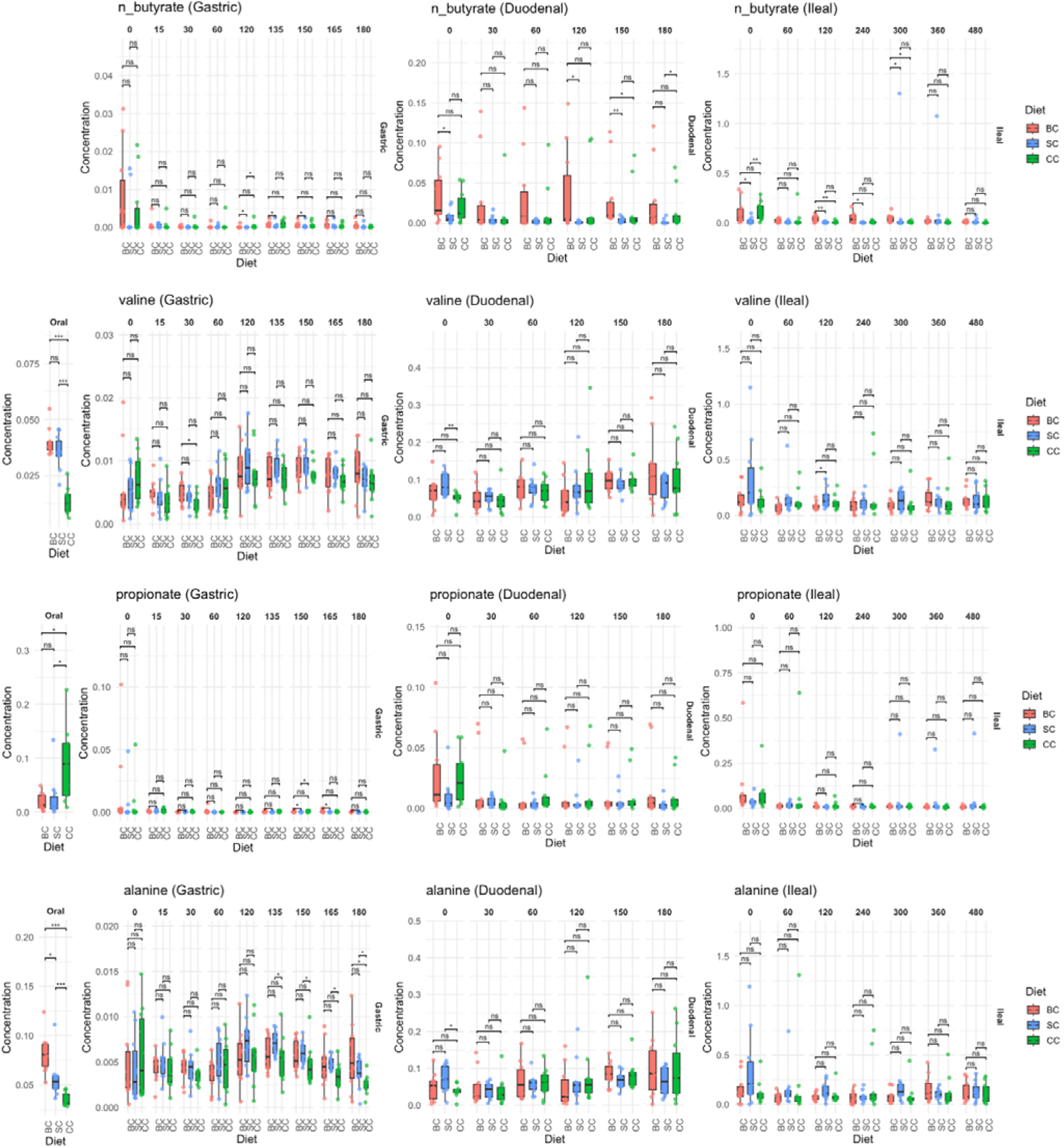
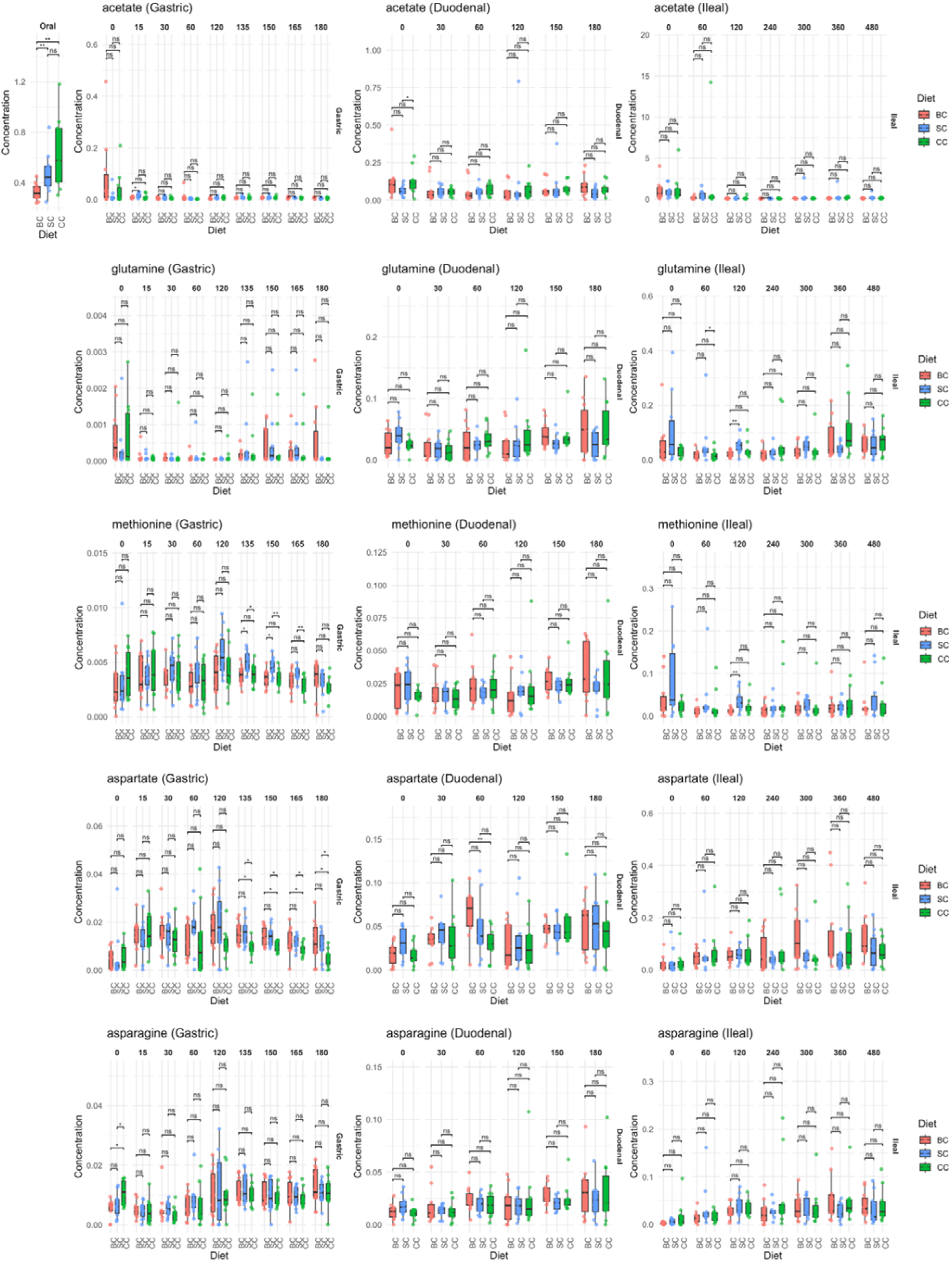
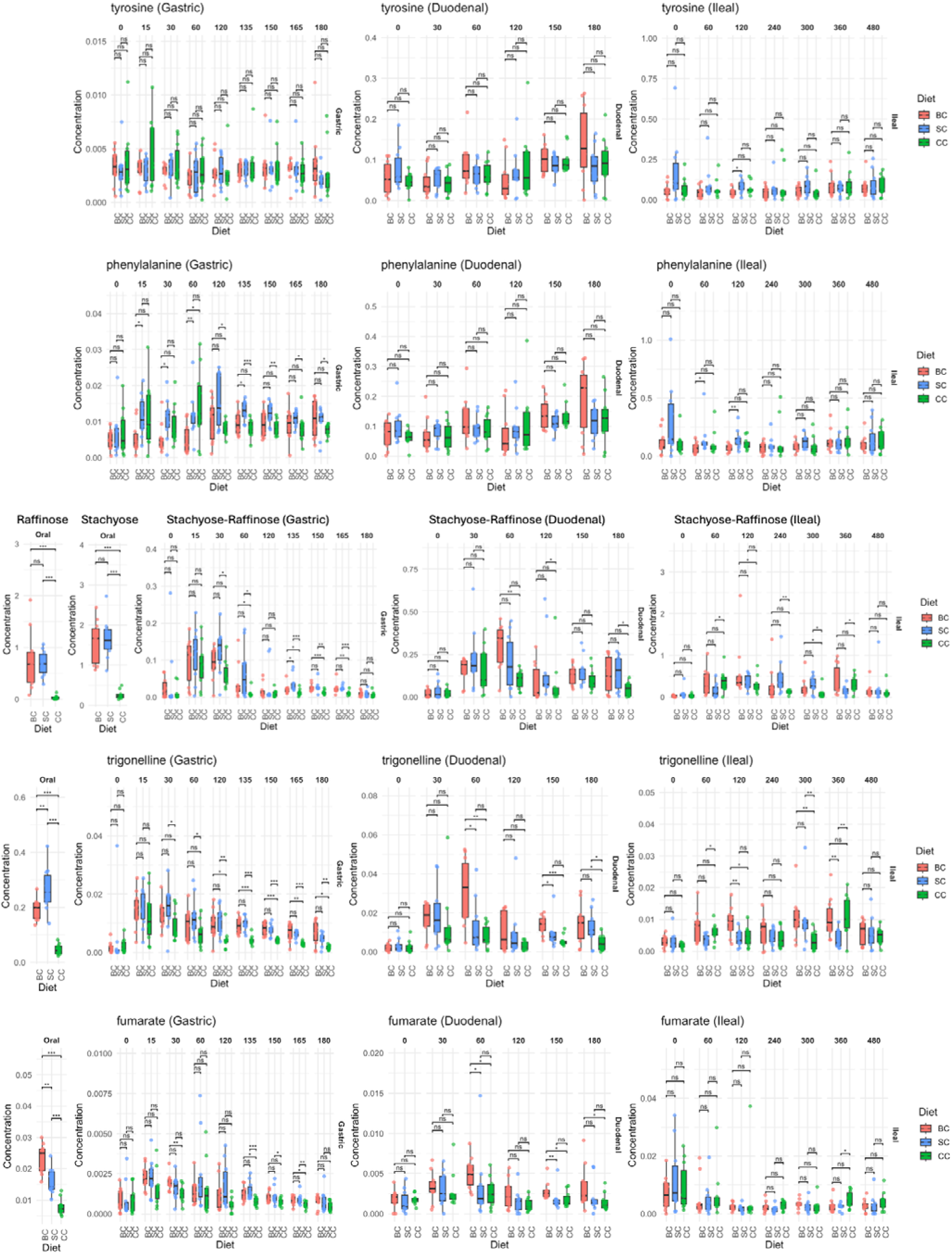
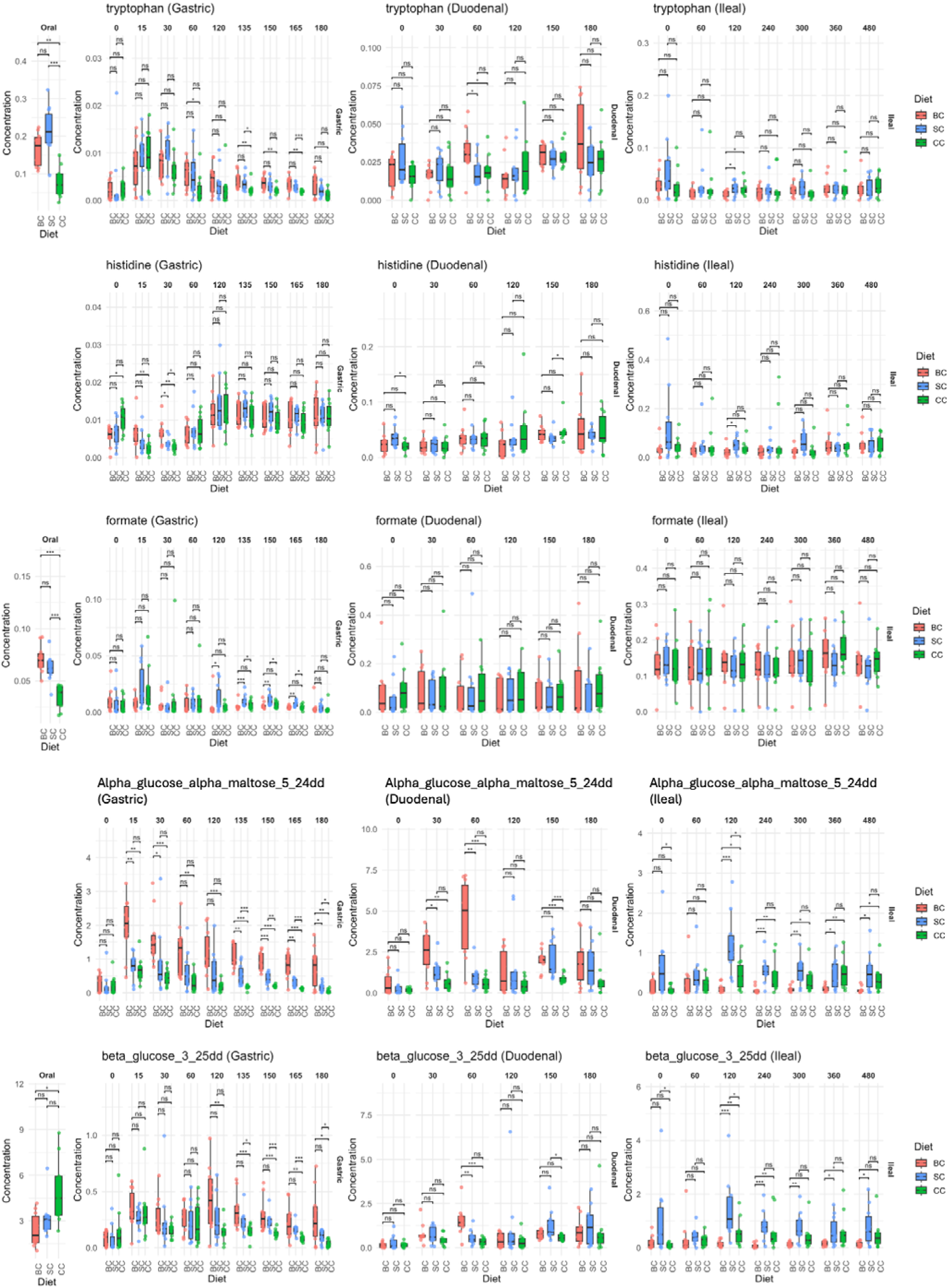
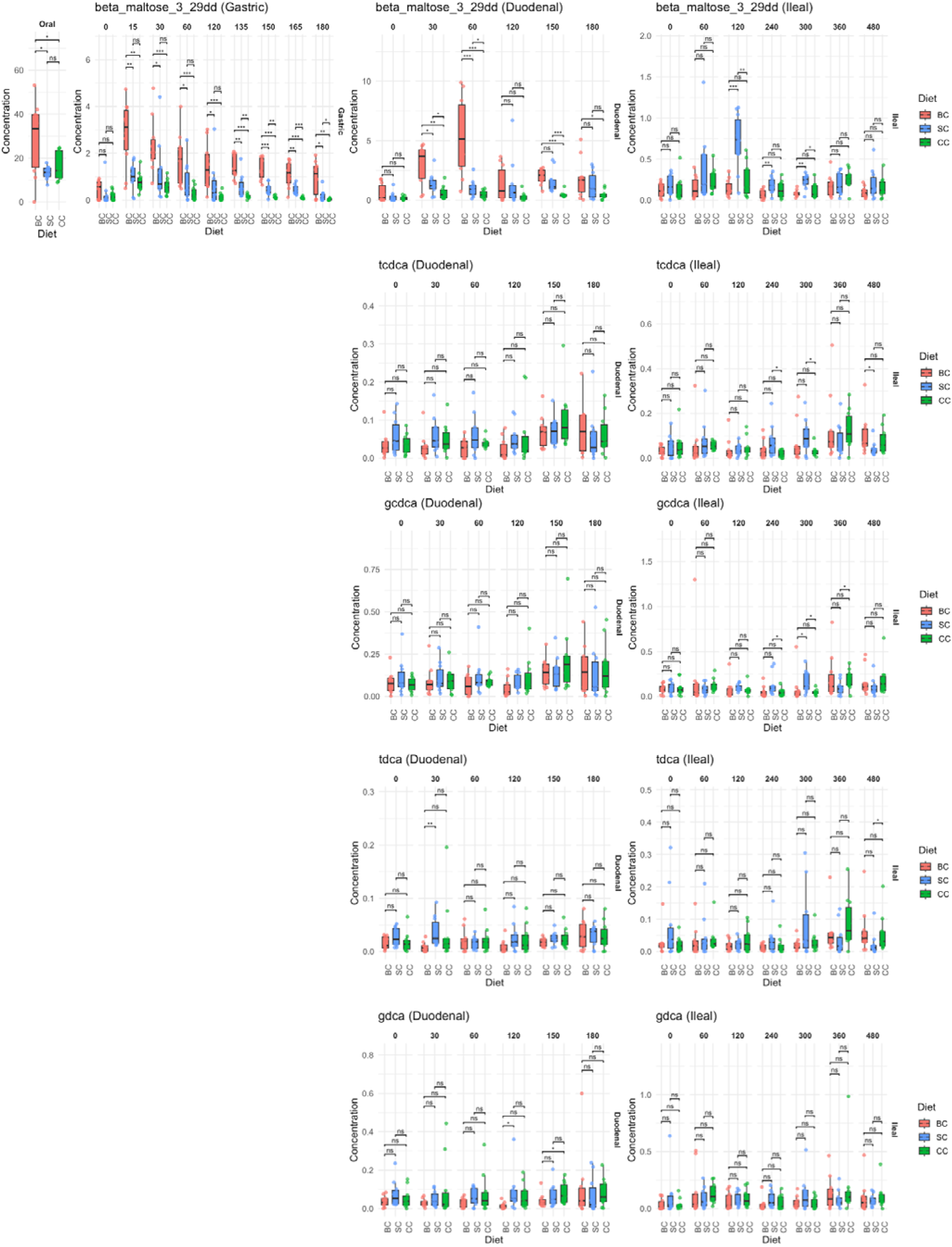
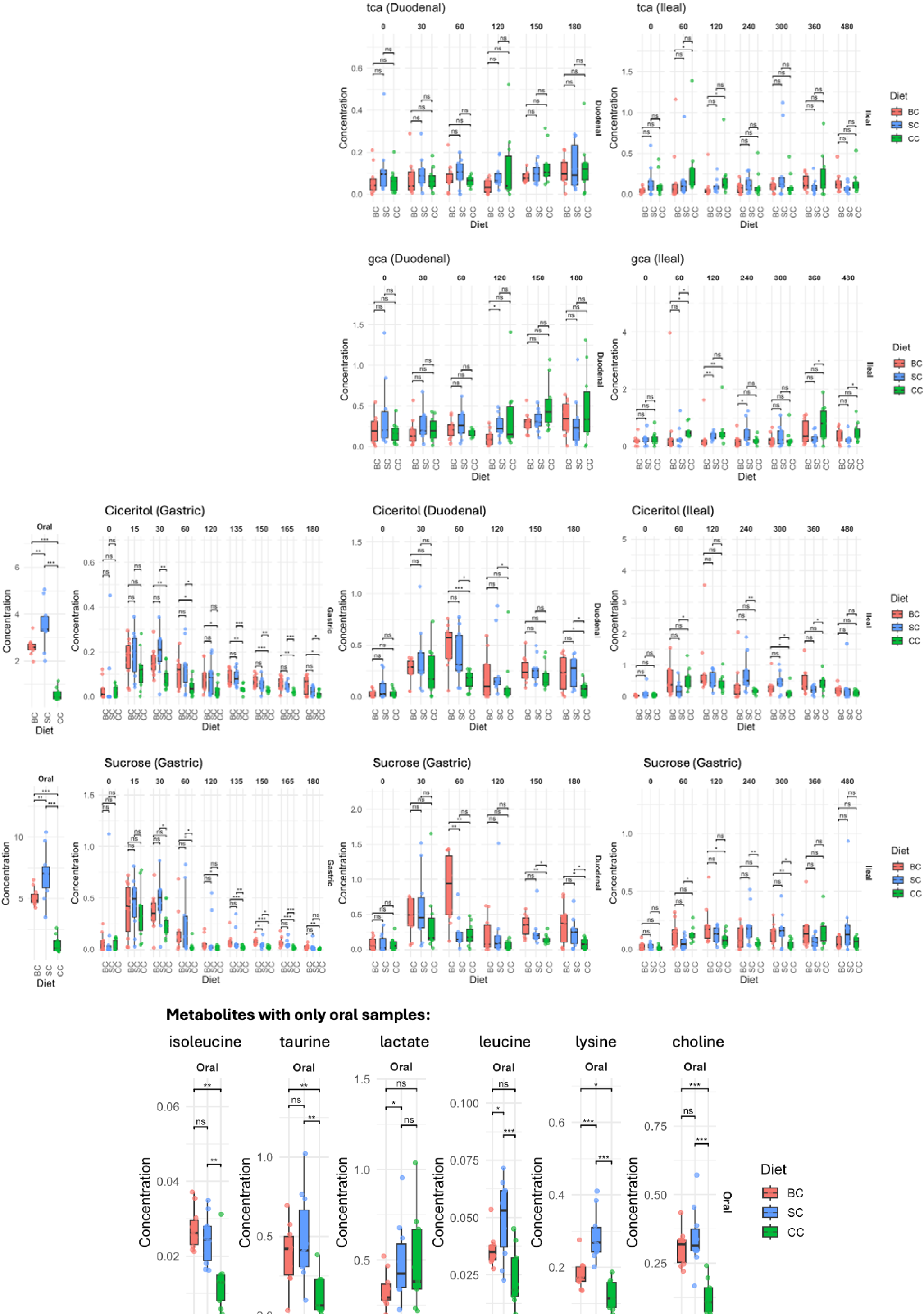
Box and whisker plots of metabolite concentrations across 3 different diets along the GI-tract, demonstrating the full range of datapoints per timepoint. Diet BC (pink) corresponds to broken cells, diet SC (blue) corresponds to single cells and diet CC (green) corresponds to cell clusters. Pairwise comparisons were assessed using the Wilcoxon rank-sum test (*** p ≤ 0.001; ** p ≤ 0.01; * p ≤ 0.05; ns, not significant).

**Extended Figure 7:**
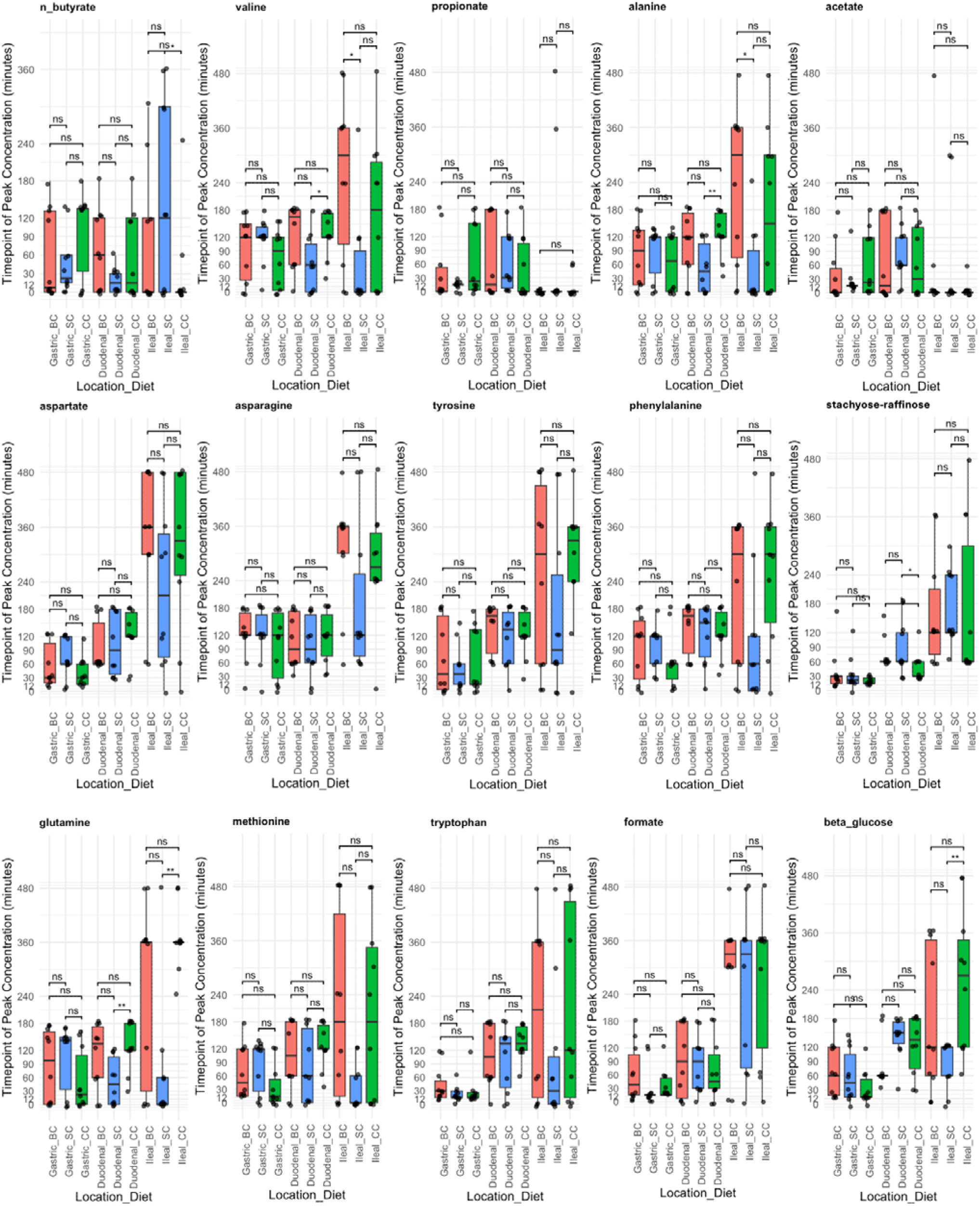
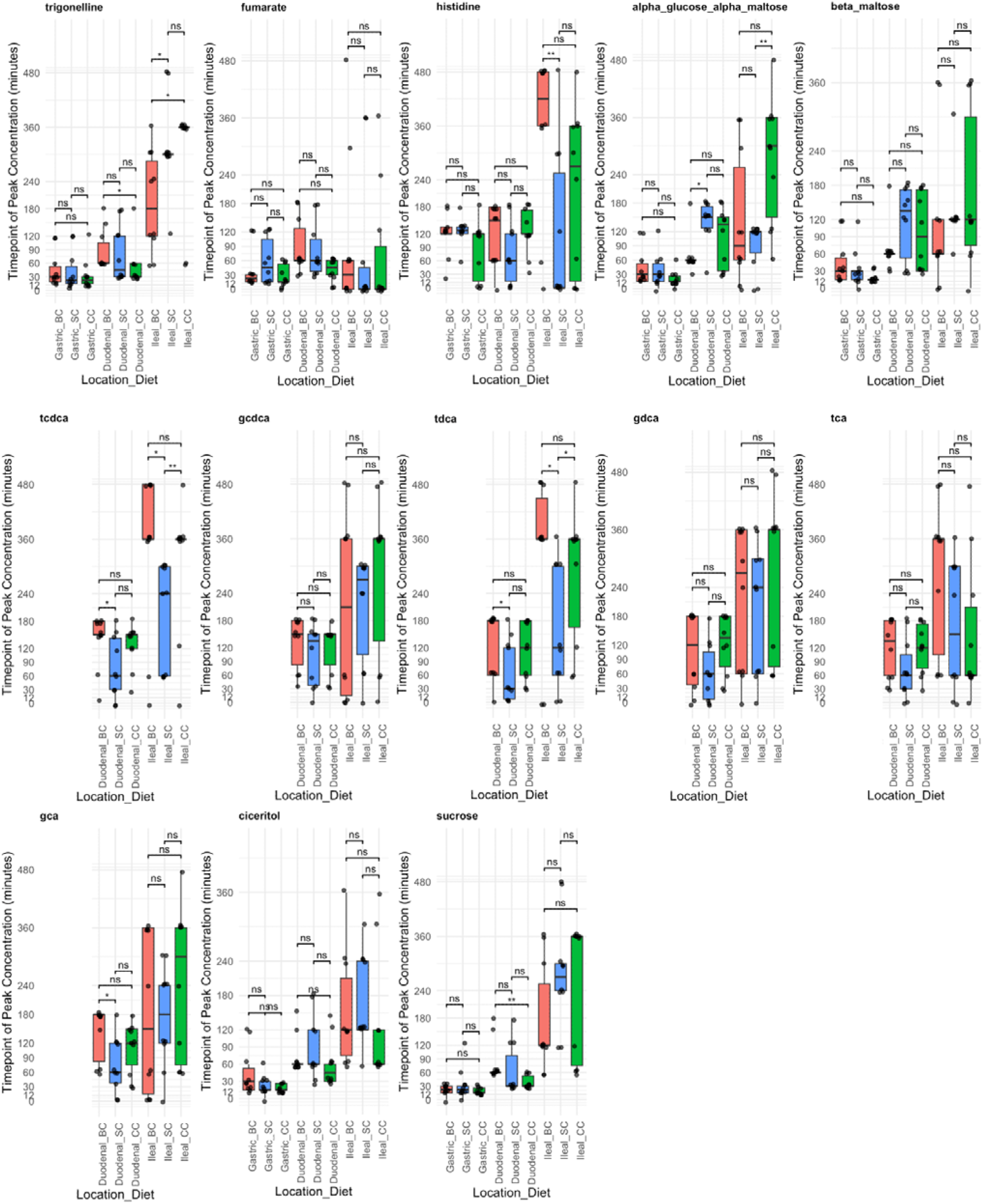
Peak concentration timepoints by diet and location. The timepoint (minutes) of maximal concentration is shown for each participant, stratified by diet [BC (broken cells), SC (single cells), CC (cell clusters)] and sampling location (gastric, duodenal, ileal). Pairwise comparisons were assessed using the Wilcoxon rank-sum test (*** p ≤ 0.001; ** p ≤ 0.01; * p ≤ 0.05; ns, not significant).

**Extended Figure 8:**
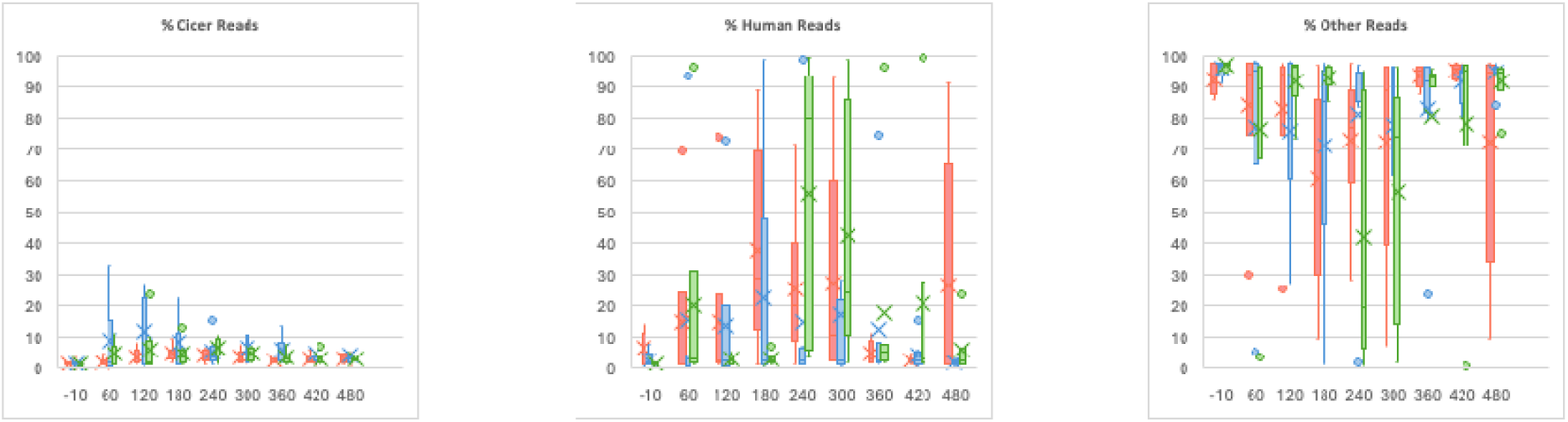
Percentage of reads from chickpea, human and other within the shotgun sequencing data. Chickpea meals legend: Broken cells (red), single cells (blue) and clustered cells (green).

**Extended Figure 9:**
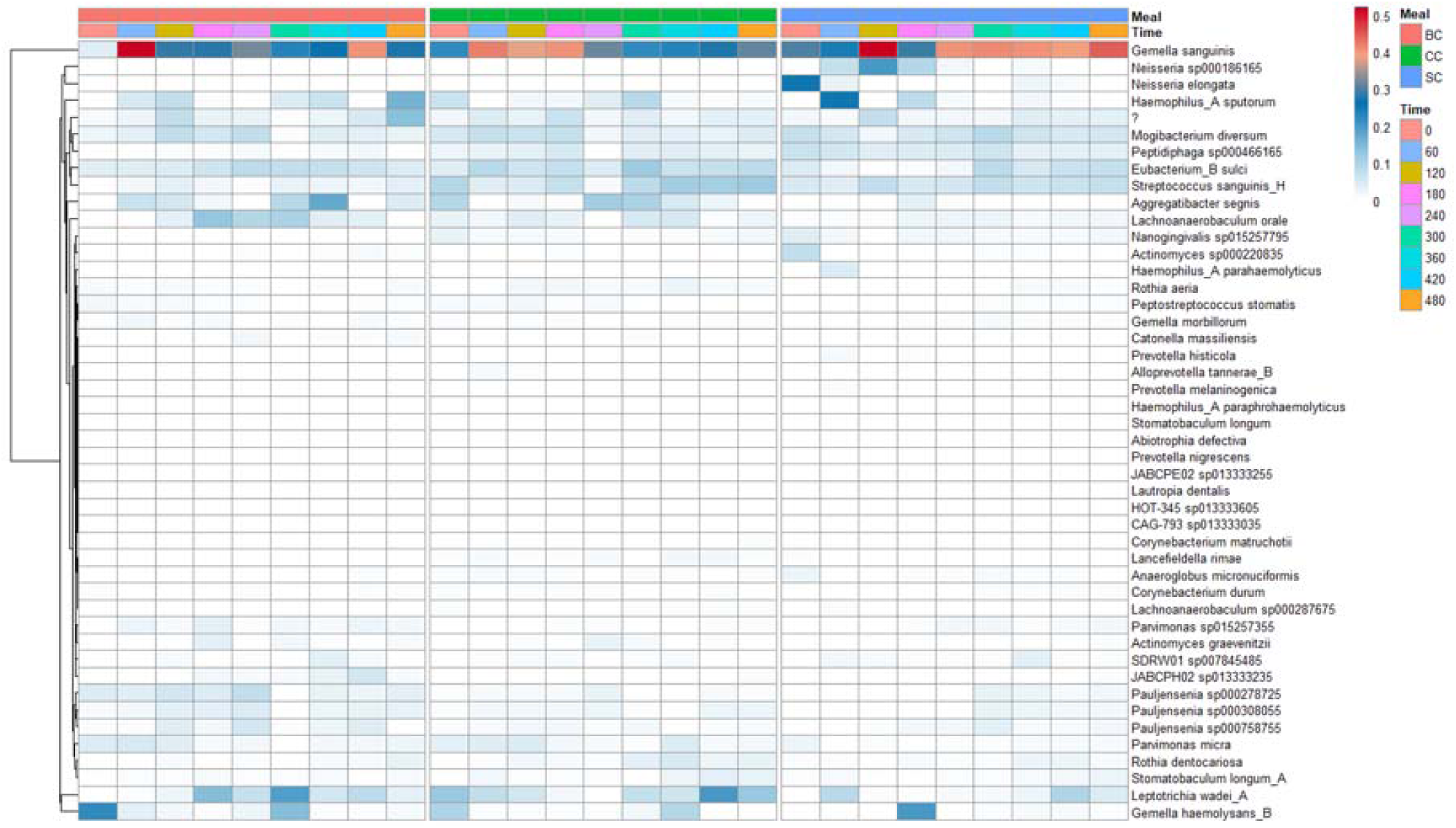
Relative abundance of microbial species in the terminal ileum. Relative abundance of microbial species in the terminal ileum shared with the oral cavity. Relative abundance of the 50 most abundant species in the terminal ileum over time during consumption of each of the 3 diets, showing only the abundance of strains shared with the oral cavity.

**Extended Figure 10:**
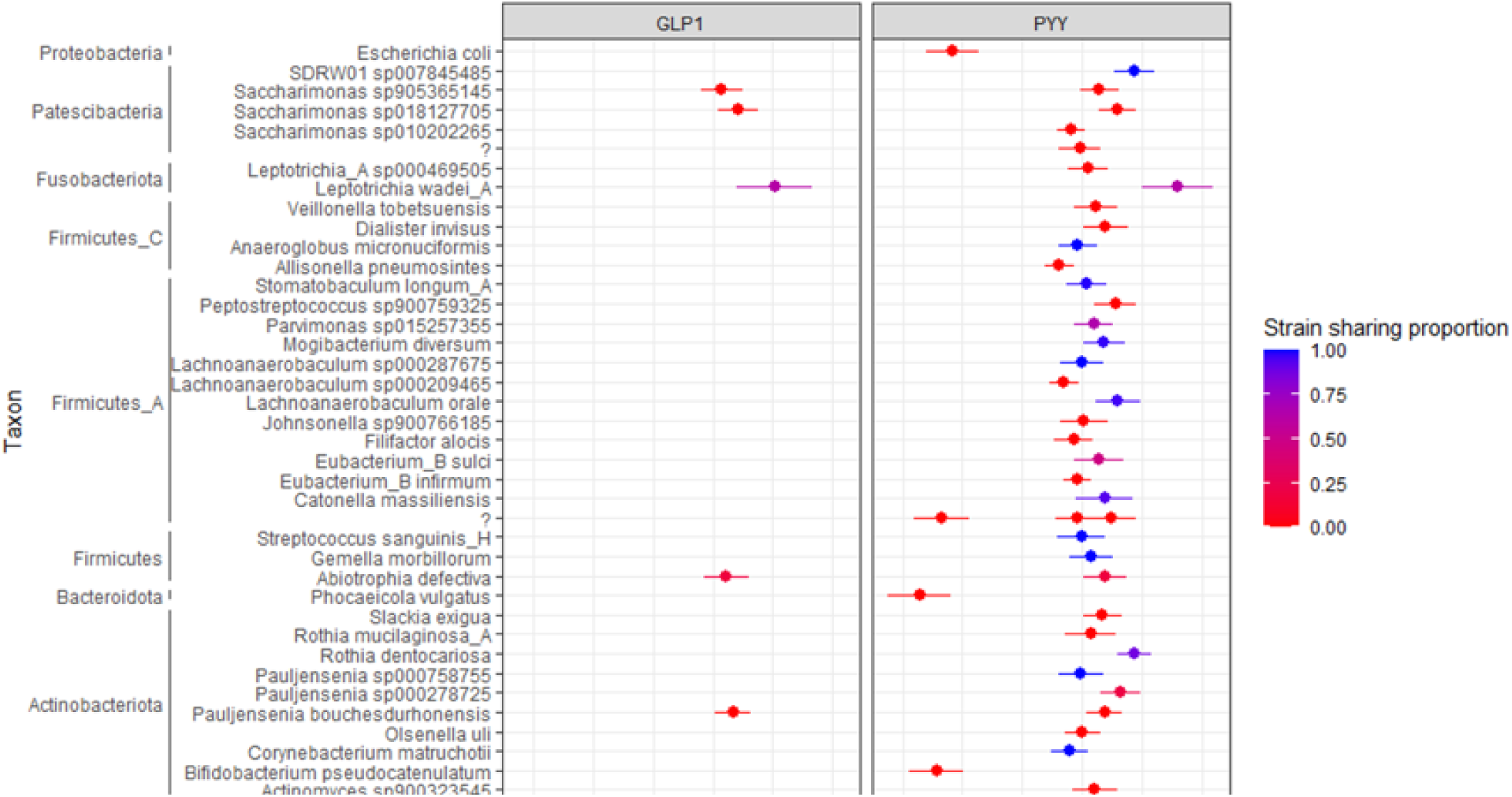
Correlations between gut hormones (GLP1 and PYY) and metagenome abundances. Associations are determined by linear modelling, with standard error shown by the error bars. Each point is coloured by the proportion of strain sharing, with red indicating that strain sharing has not been identified between the oral and ileal compartments, while blue indicates that in all instances strains are shared between oral and ileal compartments.

