## Supplementary Text 1; Supplementary Table 1-2; Supplementary Figure 1 for "Dietary Modulation of The Human Small Intestine Metabolome and Microbiome"

**Supplementary Text 1.  Meal Preparation**

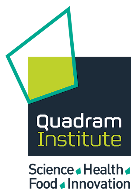

Meal Preparation

**Meals:** Chickpeas were the main ingredient of the test meals. Whole chickpeas *Cicer arietinum* L., Kabuli type, Argentine variety (7mm diameter) were supplied by AGT Poortman Ltd. These were dehulled through an abrasive process by holding 200 g batches of dry chickpea (as supplied) in a Steelbrush dehuller (Westrup pre-cleaning debranner, coarse blue roll) for 30s, then sieved on a 3mm grid to exclude fragments of testa. The resulting dehulled chickpea material was weighed out into 75.0 g portions (by a researcher independent to the study), sealed and labelled. These dehulled chickpea portions were the main ingredient for the ‘*broken cell’* (Broken) and ‘*intact separated cell’* (Intact-S) chickpea meals, which were freshly prepared on the morning of each study visit using standardised processing protocols to deliver different structures.  For the ***‘****cell cluster’* (Intact-C) meal, the dehulled chickpeas were further processed by milling to obtain intact tissue particles (1.6-2.0 mm) containing whole cells, before portioning out. These coarse particles were obtained by trickle feeding the dehulled chickpeas through a Buhler miag Vario mill set up with a Roll Gap of 1.8 mm and Roller speed 220:450 rpm on 24.5 mm diameter break rolls 5 flutes/cm break rolls in Sharp-Sharp disposition for the first break. Output material was analytically sieved to collect target size fraction (≥ 1.6 and < 2.0 mm), and the oversize material (≥ 2.0 mm) was then re-milled according to the same setting as used for the first break, but with a smaller Roll Gap (0.8 mm), to increase recovery of the target size fraction. This coarsely milled chickpea material (≥1.6 and <2.0 mm ‘semolina’) was the main ingredient for the ‘cell-cluster’ meal, and was weighed into 75 g portions in preparation for the study visits.

Each chickpea ingredient portion was then processed further, as described below,  using a Thermomix® TM5 (Vorwerk) food processor to deliver freshly cooked meals with different structures to the study participants.

For the ‘Broken cell’ and ‘Separated cells’ meal types, the pre-weighed 75 g portion of de-hulled chickpeas was rinsed with drinking water from the tap in a colander until the water ran clear, and then immersed in 1L tap water under cover overnight for ~16 h. The following morning, the soaked chickpeas were then drained, rinsed briefly with fresh tap water, and drained thoroughly before transferring to the Thermomix. For the **Broken** meal, additional tap water was then added to the soaked chickpea to achieve a total weight of 637.5 g. This mixture was then blended (forward direction, speed setting 10) for 3 min at 37 °C, which breaks the chickpea cotyledon cells, before further heating and cooking at 95 °C for 28 min, with gentle stirring (blender in reverse setting, speed setting 4). A portion (~490 g) of the cooked mixture was then weighed into a bowl and seasoned with a 115 g pot of jelly (Hartleys’ no added sugar raspberry jelly) and 15 g of sugar-free jam (Stutes no added sugar blackcurrant jam), to aid palatability, and served with drinking water (~270 mL) and served immediately to participants while still warm (above 60 °C).

The **Intact-S** meal, additional tap water was added after the overnight soak to achieve a total weight of 375.0 g, and then covered, heated and cooked at 95 °C for 80 min, with gentle stirring (blender in reverse direction, speed setting 5). A portion (~280 g) of the cooked mixture was then weighed into a bowl, seasoned as described above, and served immediately while still warm, together with drinking water (~480 mL).

For the **Intact-C** meal, the soaked semolina were drained and transferred into the Thermomix together with additional tap water to achieve a total wet weight of 300 g. This was then cooked for 50 min at 95 °C with gentle stirring (reverse, speed setting 1). A serving of ~120 g drained cooked chickpea solids was then transferred into a bowl, seasoned as above, and served immediately while warm together with drinking water.

For all meals, the volume of water served in the drinking glass was adjusted based on the cooked meal weight, so that all meals provided 30 g total starch from chickpea, the same amount of seasoning and similar total mass (~760 g).

Supplementary Table 1. Inclusion and exclusion criteria for participants

| **Inclusion Criteria** | **Exclusion Criteria** |
| --- | --- |
| Male or female | Abnormal ECG |
| Age between 18-65 years (inclusive) | Screening blood results outside of normal reference values.  Fasting glucose 3.9-5.6 mmol/L  Haemoglobin M125-170/ F114-150 g/L  White blood cell count 4.2-11.2 x 10^9^/L  Red blood cell count 3.73-4.96 million/mcL |
| Body mass index (BMI) of 18-30 kg/m^2^ | Weight change of ≥ 5kg in the preceding 2 months |
| Willingness and ability to give written informed consent and willingness and ability to understand, to participate and to comply with the study requirements | Current Smokers |
|  | History of substance abuse and/or excess alcohol intake |
|  | Cancer |
|  | Gastrointestinal disease e.g., inflammatory bowel disease or irritable bowel syndrome, kidney disease, liver disease and pancreatitis |
|  | Pregnancy |
|  | Diabetes or cardiovascular disease |
|  | Participant in a research study or blood donation 12 weeks prior |
|  | Started new medication within the last 3 months likely to interfere with glucose and energy metabolism, appetite regulation and hormonal balance, including: anti-inflammatory drugs or steroids, antibiotics, androgens, phenytoin, erythromycin, or thyroid hormones. |

Supplementary Table 2 Baseline characteristics for participants

| **Variable** | **Baseline measurements** |
| --- | --- |
| Gender (M:F) | 6:4 |
| Age (years) | 30.8 ± 2.41^1^ |
| Height (m) | 1.70 ± 0.02^1^ |
| Weight (kg) | 72.07 ± 3.20^1^ |
| BMI (kg/m^2^) | 24.91 ± 0.82^1^ |
| Body Fat (%) | 17.35 ± 1.91^1^ |

^1^Results presented as Mean ± SEM

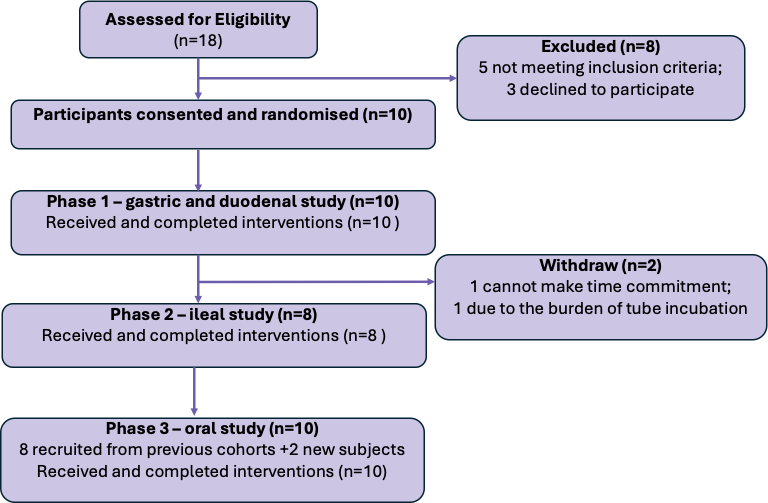

Supplementary Figure 1. CONSORT flow diagram illustrating the study procedures.
