## Supplementary material for "Dietary Modulation of The Human Small Intestine Metabolome and Microbiome": Study protocol

**Determining the effect of chickpea tissue-structures on metabolic responses, satiety regulation and gut content along the entire gastrointestinal tract**

Main Sponsor	Imperial College London

REC Reference number: 19/LO/0950

**Protocol authorised by**

**Professor Gary Frost,**

Department of Investigative Medicine,

Hammersmith Hospital Campus, Imperial College,

6^th^ Floor, Commonwealth Building,

Du Cane Road, London,

W12 0NN

**Dr. Dominic Blunt**

Imaging Department, Charing Cross Hospital

Fulham Palace Road, London

W6 8RF

**Dr. Cathrina Edwards**

Quadram Institute Bioscience,

Norwich Research Park

NR4 7UQ

**Dr. Martina Tashkova**

Department of Investigative Medicine,

Hammersmith Hospital Campus, Imperial College,

7^th^ Floor, Commonwealth Building,

Du Cane Road, London,

W12 0NN

**Dr. Claire Byrne**

Department of Investigative Medicine,

Hammersmith Hospital Campus, Imperial College,

7^th^ Floor, Commonwealth Building,

Du Cane Road, London,

W12 0NN

**Dr Katerina Petropoulou**

Department of Investigative Medicine,

Hammersmith Hospital Campus, Imperial College,

7^th^ Floor, Commonwealth Building,

Du Cane Road, London,

W12 0NN

**Dr Mingzhu Cai**

Department of Investigative Medicine,

Hammersmith Hospital Campus, Imperial College,

7^th^ Floor, Commonwealth Building,

Du Cane Road, London,

W12 0NN

Long title:

**Determining the effect of chickpea tissue-structures on appetite, satiety and gut content along the entire GI tract.**

Short title:

**Chickpea tissue-structures and gut hormone response.**

**Study Management Group**

Chief Investigator	Professor Gary Frost

Co-investigators

Dr. Dominic Blunt

Dr. Cathrina Edwards

Dr. Martina Tashkova

Dr. Claire Byrne

Dr. Katerina Petropoulou

Dr. Mingzhu Cai

**Sponsor**

Imperial College is the main research sponsor for this study. For further information regarding the sponsorship conditions, please contact the Research Governance Manager at:

Joint Research Compliance Office

Imperial College London and Imperial College Healthcare NHS Trust

Room 215, Level 2, Medical School Building

Norfolk Place

London, W2 1PG

[www.imperial.ac.uk/clinicalresearchoffice](http://www.imperial.ac.uk/clinicalresearchoffice)

**PROBLEMS RELATED TO THIS TRIAL SHOULD BE REFERRED TO PROFESSOR GARY FROST**

1. INTRODUCTION
2. STUDY METHODOLOGY
   2.1. STUDY DESIGN

2.1.1.	Study Visit 1 – UPPER GI

2.1.2.	Study Visit 2,3,4 – LOWER GI

2.1.3. Study Visit 5- ORAL DIGESTION

2.2. INCLUSION/ EXCLUSION CRITERIA

3. WITHDRAWL CRITERIA AND ADVERSE EVENTS
   3.1. WITHDRAWL CRITERIA
   3.2. ADVERSE EVENTS
4. REPORTING PROCEDURES
   4.1. NON-SERIOUS AEs
   4.2. SERIOUS AEs
5. REGULATORY ISSUES

5.1. ETHICS APPROVAL
5.2. CONFIDENTIALITY
5.3. INDEMNITY
5.4. SPONSOR
5.5. FUNDING
5.6. REIMBURSMENT AND CONCENT
5.7. AUDITS AND INSPECTIONS

6. PUBLICATION POLICY
7. REFERENCES

**1. INTRODUCTION**

A number of reports globally demonstrate the rates of obesity and type 2 diabetes continue to increase. It has been estimated that more than 50% of the adult population are overweight (Wang, 2011) and that one in 17 people has either diagnosed or undiagnosed diabetes in the UK (Diabetes, U. K, 2014). These statistics highlight the importance of the maintenance of a energy balance and glucose homeostasis.

Chickpeas are an excellent source of high-quality protein and dietary fibre. Previous research has reported that chickpea starch is more resistant to digestion in the small intestine, which associates with lower bioavailability of glucose and improved bowel health (Nestel, 2004). It has been demonstrated that a controlled diet with chickpeas results in decreases in plasma glucose and insulin concentration (Nestel, 2004), as well as reductions in serum total cholesterol (TC) and low-density lipoprotein-cholesterol (LDL-C) (Pittaway, 2007), potentially contributing to lower risk of type 2 diabetes and coronary heart disease (CHD). Furthermore, compared to a wheat-based meal, greater satiety was reported by some participants (Pittaway, 2007; Zafar, 2017), possibly leading to lower energy intake and improved weight control. However, there has been limited research into chickpea and its potential health benefits and the stimulation of gut hormone secretion. Chickpeas are high in resistant starches and protein, and these nutrients have been shown to stimulate gut hormone secretion, including gastric-inhibitory peptide (GIP), glucagon-like peptide-1 (GLP-1) and peptide YY (PYY) that could regulate glucose homeostasis (Raben, 1994; Zhou, 2008; Smeets, 2008). These suggest that further research is needed to investigate the relationship between chickpeas supplementation and gut hormone secretion.

The microstructure of plant food, which is altered during processing such as grinding, heating or fermentation that could break cellular structure and thereby influence Type 1 Resistant Starch content, has been shown to affect nutrient bioavailability and digestion process (Sensory, 2014). Thus, this project will improve understanding of the relationship among food structure ranging in processing, nutrient bioavailability and chickpea-induced release of PYY, GIP and GLP-1.

The present study will combine two different methodologies for sampling from the gastrointestinal tract, which have been used in previous studies by our research group (REC references: 17/LO/0354, 15/LO/0184).

Overall, the aim of this study is to investigate the impact of different chickpea tissue-structures on gut hormone secretion, thus explaining the chickpeas’ influences on glucose control and satiety reported in previous studies.

1. **STUDY METHODOLOGY**

Study visits 1-4

**Participants:** 15 healthy male and female volunteers. This is a pilot study in a new area and therefore a formal power calculation is not possible.

**Recruitment:**

Participants will be recruited from existing healthy volunteer databases and by advertisement in public places. Adverts will be placed in newspapers and put up in public buildings. A contact number on the advert will enable potential participants to contact the research team at Imperial College London. Participation in the study will be entirely voluntary. No undue influence will be exerted by the researchers. Participants will be free to withdraw from the study at any time.

Once potential participants have responded to a study advertisement a researcher will arrange a short telephone interview to explain the study. Following this a more formal interview will be arranged in order for the researcher to determine if the potential participant meets the inclusion and exclusion criteria. This also gives the potential participants a chance to ask any questions they may have about the study. Prior to the interview they will have been sent a written participant information sheet by email or by post.

Study visit 5

**Recruitment target**: The recruiting target for study visit 5 is 10 participants. This number has been determined to provide sufficient samples for analyses.

**Participant identification and recruitment process:**

(1) **From participants who completed study visit 1-4**

Potential participants will be initially identified from the pool of previous study subjects who have completed study visits 1-4. We will assess the eligibility of these potential participants based on the following criteria:

- Consent status: we will verify whether the subject has consented to be contacted for further visits. This will be conducted by the researchers who initially obtained the consent.
- Registration with healthy volunteer dataset: we will confirm whether the subject is still listed in the healthy volunteer database. This will be carried out by administrators from NIHR Imperial Clinical Research Facility who have access to the database.

Contacting eligible participants： only participants who have given consent to be contacted and are currently registered as healthy volunteers will receive an email invitation to participant in the study visit 5. Emails will be sent by a member from the research team as soon as the amendment is approved.

Accepting of participants: all eligible participants who express willingness to take part in the study visit 5 will be accepted, even if the number exceeds the target of 10.

(2) **Recruitment for new participants**

If fewer than 10 former participants are available and willing to participate, new participants will be recruited from the healthy volunteer database. These participants will take part in study visit 5 only.

All participants, whether former or new, will undergo a thorough informed consent process before participating in study visit 5.

**2.1.	STUDY DESIGN**

**This study includes five study visits at clinical research facility**

This will follows a randomized crossover design involving five study visits. Participants will be clinical research facility inpatients for four days on study visit 1, and 3 days on study visit 2,3,4. There will be at least 7 days between visits 2-3 and 3-4.

The study visit 5 will last for three hours at clinical research facility.

**Health Screening:**

Participants will attend the NIHR/Wellcome Trust Imperial Clinical Research Facility at Hammersmith Hospital where their eligibility will be assessed. A pre-screening questionnaire will be done to record the personal information, body composition and medical history.They will have a blood test (HbA1c, FBC, LFT, U&E and lipids) and height and weight measurements will also be taken. They will also have an electrocardiogram (ECG) and blood pressure will be recorded. All women of child bearing age will have a pregnancy test.

***Study Visit 1***

The day prior to the study visit, the participants will be requested to refrain from strenuous exercise and alcohol and to arrive having fasted from the evening before. The following morning participants will be asked to attend the NIHR Imperial Clinical Research Facility (CRF) at Hammersmith Hospital for 4 days (3 nights).

**Day 1**

Enteral feeding tubes will be placed to allow for sampling of intestinal content from the stomach and small intestine throughout the study visit. The enteral feeding tubes will be placed by a trained medical professional in the Imperial Clinical Research Facility using the CORPAK feeding tubes that track the position of the tube during placement without the need for x-rays. This system has been used in previous studies by our research group (REC Ref: 15/LO/0184). These tubes will remain in place for the duration of the 4 day visit. The diet of all participants will be standardised throughout the visit.

**Day 2-4**

On the morning of Day 2, an intravenous cannula will be inserted to allow for blood sampling and will remain in place for the remainder of the study visit.

Each morning, two fasting blood samples and gastric and duodenal samples will be taken prior to the test meal. In addition, two baseline subjective appetite measures, assessed by visual analogue scale, and two baseline breath hydrogen measurements will be collected in real-time using a gastrolyser.

Each day, in a random order, participants will then receive a standardised breakfast made from cooked chickpeas with different structures:

1. Broken cells from chickpeas (control)

2. Individual cells from chickpeas

3. Clusters of cells from chickpeas

The control breakfast will provide the same total starch content as the test interventions. The chickpea material will be provided by Quadram Institute Bioscience.The recipe of the breakfast will be designed by the research group from Imperial College London and Quadram Institute Bioscience. The food material will be purchased from Sainsbury's or another UK food supermarket. Volunteers will be randomized using the 'sealed envelope' website.

Further blood samples will be collected at 15, 30, 45, 60, 90, 120, 150, 180 min following the test meal in order to measure hormones, metabolites and inflammatory signals. 10 ml of blood will be taken at each blood sample. 300 ml of blood will be taken during the 4 day study visit.

Gastric and duodenal content will be taken at 15, 30, 45, 60, 75, 90, 105, 120, 135, 150, 165, 180 min. Microscopy and metabolomics assessment will be performed to assess the impact of initial digestion on the breakdown of the different chickpea structures.

Subjective appetite measures will be collected at 15, 30, 45, 60, 90, 120, 150, 180 min using VAS questionnaire. Breath hydrogen will be measured at 0, 60, 120, 180 min.

Urine will be collected throughout the study visit to measure metabolite concentrations. Subjects will empty their bladder before the test breakfast and collect all urine thereafter for a period of 180 min. Subjects will be provided with an appropriate measurement container to collect urine.

Following the 180 min sample on the fourth study day, the enteral tubes and intravenous cannula will be removed and volunteers will be discharged from the Clinical Research Facility.

A lunchtime meal (at 4 h) will be provided in surplus, and dietary intakes recorded such that the participant's ad libitum energy intake can be determined. A fixed dinner-time meal will be provided at 9 h. Water (for the first 4 h) and beverages are provided throughout the day.

After study visits, gastric and duodenal samples will be sent to Quadram Institute Bioscience and stored for subsequent analysis.

***Study Visits 2,3,4***

Study Visits 2,3, and 4 are focussed on events occurring in the lower small intestine, and the microbial and metabolic responses to these events.

**Day 1**

The day prior to each 3-day study visit, participants will be requested to refrain from strenuous exercise, caffeine and alcohol. Participants will then be requested to fast overnight (they are allowed to drink water) and will come to the Imaging Department at Charing Cross Hospital for the insertion of nasoenteric tubes. Following the tube insertion, participants will return to the NIHR Imperial CRF to complete the three day study. They will return to the CRF at Hammersmith Hospital in a taxi and will be accompanied by a member of the study team.

On day 1 of the study, all female participants of child bearing age will be asked to provide a urine specimen in order to perform a pregnancy test prior to placement of the tube. A nasoenteric tube will then be inserted through the nose, with a small balloon at the terminal end which is inflated and used to carry the tube through the small intestine by peristalsis. Once the tube reaches the terminal ileum, the balloon is deflated and the tube is restrained from additional movement for the rest of the 3-day study visit. The tube position will be confirmed by fluoroscopy and administration of diluted barium sulphate. The tubes will be inserted at Charing Cross Hospital following a procedure which has been used in previous studies by our research group (17/LO/0354). Following the tube insertion, participants will travel in a taxi accompanied by a member of the study team to the NIHR/Wellcome Trust Imperial CRF to complete the four day study visit.

During these 3 separate study visits, volunteers will be provided with chickpea-enriched diets differing in which the structure of the chickpea component differs. The chickpea material will be provided by Quadram Institute Bioscience. The recipe of the diet will be designed by the research group from Imperial College London and Quadram Institute Bioscience. The food material will be purchased from Sainsbury's or another UK food supermarket Each visit, in a randomised order, volunteers will receive a standardised low-fibre background diet supplemented with one of the following:

1. broken cells from chickpeas (control)

2. individual cells from chickpeas

3. chickpea cell clusters

Volunteers will be fed one of the diets over the 3-day study period. The diet will start following the tube placement on Day 1 and end on Day 3. Volunteers will also be asked to collect a stool sample on each day of the 3-day study period.

Volunteers will be randomized using the 'sealed envelope' website.

Ileal sample collection will start **on day 2**, with two baseline samples taken before breakfast and 60, 120, 180, 240, 300, 360, 420 and 480 min after breakfast for metabolomic analysis, which will include 1H NMR spectroscopy, ultra-performance LC-MS and GC-MS, microbiological analysis and for microscopy.

**On day 3**, ileal samples will be collected as described on day 2 and will be matched with blood sampling and visual analogue scales (VAS) to assess appetite responses. An intravenous cannula will be inserted on the morning of day 3 and two fasting blood samples will be collected before breakfast. After breakfast, blood samples will be collected at 60, 120, 180, 240, 300, 360, 420 and 480 min to measure hormones and metabolites. 100 ml (10 × 10 ml) will be collected throughout each study visit. At each time point, volunteers will be asked to complete VAS to assess subjective feelings of hunger, fullness and nausea. Breath hydrogen concentrations will also be measured at the same time.

At the end of day 3, the cannula and the nasoenteric tube will be removed. It is expected that the majority of participants will have the naso-enteric tubes removed through the nose at the CRF Unit. If more than mild discomfort seems likely to be caused by the removal of the tube, they will be transported to Charing Cross Hospital in a taxi accompanied by a trained medical professional and will have the tube removed under fluoroscopy, or the nasal end of the tube will be cut and the rest of the tube will be allowed to pass rectally.

Collected tissue samples stored may be used in future ethically approved studies.

After study visits, ileum and stool samples will be sent to Quadram Institute Bioscience and stored for subsequent analysis.

***Study Visit 5***

The study will investigate the oral digestion events. Participants will be instructed to chew portions of chickpea meals (as listed below, consistent with study visit 1-4) in a randomised order and split it out.

1. Broken cells from chickpeas (control)

2. Individual cells from chickpeas

3. Clusters of cells from chickpeas

The post-chewing food samples will be collected to analyse nutrient release and the breakdown of structure during oral digestion. These findings, combined with our previous investigations into gastric and intestinal digestion, will provide us an enhanced understanding of how food structure affects human digestion and metabolism.

**2.2.	INCLUSION/ EXCLUSION CRITERIA**

*Inclusion criteria:*

• Male or female

• Age between 18-65 years (inclusive)

• Body mass index (BMI) of 18-30	kg/m2

• Willingness and ability to give written informed consent and willingness and ability to understand, to participate and to comply with the study requirements

*Exclusion criteria:*

• Abnormal ECG

• Screening blood results outside of normal reference values

• Weight change of ≥ 5kg in the preceding 2 months

• Current smokers

• History of substance abuse and/or excess alcohol intake • Pregnancy • Diabetes • Cardiovascular disease

• Cancer

• Gastrointestinal disease e.g. inflammatory bowel disease or irritable bowel syndrome

• Kidney disease

• Liver disease

• Pancreatitis

• Started new medication within the last 3 months likely to interfere with energy metabolism, appetite regulation and hormonal balance, including: anti-inflammatory drugs or steroids, antibiotics, androgens, phenytoin, erythromycin or thyroid hormones.

• Participation in a research study in the 12 week period prior to entering this study.

• Any blood donation within the 12 week period prior to entering this study

Any participants with the above conditions would already have an altered pattern of hormones and inflammatory molecules because of their disease process and would therefore give us confounding or misleading results.

Support of number of volunteers: This is a pilot study in a new area and therefore a power calculation is not possible.

3. **WITHDRAWL CRITERIA AND ADVERSE EVENTS**
   3.1. **WITHDRAWAL CRITERIA**

The safety of the study participants takes priority. Any significant adverse event (as assessed by the researchers) will halt the study and the ethics committee and sponsor will be informed as per standard protocol. All adverse events will be recorded and investigators will review each adverse event as it arises. In addition, participants will be free to withdraw at any time and are not required to give a reason.

3.2. **ADVERSE EVENTS**

**Adverse Event (AE):** Any untoward medical occurrence in a patient or clinical study subject.

**Serious Adverse Event (SAE):** Any untoward and unexpected medical occurrence that:

- results in death
- is life- threatening – refers to an event in which the subject was at risk of death at the time of the event; it does not refer to an event which hypothetically might have caused death if it was more severe.
- requires hospitalisation, or prolongation of existing inpatients’ hospitalisation.
- results in persistent or significant disability or incapacity
- is a congenital abnormality or birth defect

Medical judgement should be exercised in deciding whether an AE is serious in other situations. Important AEs that are not immediately life threatening or do not result in death or hospitalisation but may jeopardise the subject or may require intervention to prevent one of the other outcomes listed in the definition above, should also be considered serious.

4. **REPORTING PROCEDURES**

All adverse events should be reported. Depending on the nature of the event the reporting procedures below should be followed. Any questions concerning adverse event reporting should be directed to the Chief Investigator in the first instance.

4.1. **Non-serious AEs**

All such events, whether expected or not, should be recorded.

4.2. **Serious AEs (SEAs)**

An SAE form should be completed and emailed to the Chief Investigator within 24 h. However, relapse, death and hospitalisations for elective treatment of a pre-existing condition do not need reporting as SAEs.

All SAEs should be reported to the XXXX Research Ethics Committee where in the opinion of the Chief Investigator the event was:

- ‘related’, i.e. resulted from the administration of any of the research procedures; and
- ‘unexpected’, i.e. an event that is not listed in the protocol as an expected occurrence.

Reports of related and unexpected SAEs should be submitted within 15 days of the Chief Investigator becoming aware of the event, using the NRES SAE form.

Local investigators should report any SAEs to the sponsor and their Local Research Ethics Committee and/ or Research and Development Office.

**Contact details for reporting SAEs**

**, attention Professor Gary Frost**

**Please send SAE forms to Professor Gary Frost**

5. **REGULATORY ISSUES**
   5.1. **ETHICS APPROVAL**

The Chief Investigator has obtained approval from the HRA and Research Ethics Committee.  The study must also receive confirmation of capacity and capability from each participating NHS Trust before accepting participants into the study. The study will be conducted in accordance with the recommendations for physicians involved in research on human subjects adopted by the 18th World Medical Assembly, Helsinki 1964 and later revisions

5.2. **CONFIDENTIALITY**

The Chief Investigator will preserve the confidentiality of participants in the study and is registered under the Data Protection Act. The Principal Investigator will preserve the confidentiality of participants taking part in the study and is registered under the Data Protection Act 2018. Signed consent forms will be kept in a locked filing cabinet in a locked office in the Section of Investigative Medicine, Imperial College London. These forms will contain the participant names and an individual study code. All other data will contain the individual study code and no other participant identifying information. This will make data anonymous.

Anonymised data will securely forward to researchers in the Food Innovation Health Programme at Quadram Institute Bioscience,located at Norwich Research Park, Colney Ln, Norwich NR4 7UA, after a non-disclosure agreement specific to this study is signed

5.3. **INDEMNITY**

Imperial College holds negligent harm and non-negligent harm insurance policies, which apply to this study.

5.4. **SPONSOR**

Imperial College London will act as the main sponsor for this study. Delegated responsibilities will be assigned to the NHS trusts taking part in this study.

5.5. **FUNDING**

The study is funded by a BBSRC Strategic Programme Grant (BB/R012512/1) awarded to the Quadram Institute Bioscience (QIB). Prof. Gary Frost is a Principal Investigator within the QIB Food Innovation and Health Programme (lead PI: Prof. Richard Mithen) and has been allocated funds (£688,179) for completion of this study.

5.6. **REIMBURSMENT AND CONSENT**

Recruitment posters will be placed on Imperial College London campuses and Imperial College Healthcare NHS Trust Sites (South Kensington, Hammersmith Hospital, Charing Cross,St. Mary’s and Queen Charlotte’s Hospital).  Adverts may also be placed in newspapers or magazines, or on the Imperial College Healthcare NHS Trust Internet Homepage and will use the same text as that used on the recruitment poster, although layout may vary.

Written and informed consent will be taken by a member of the research team who has experience in obtaining informed consent. Those participants who agree to take part will be asked to sign a consent form before any study procedure is started. Participation in the study will be entirely voluntary. No undue influence will be exerted by the researchers. Participants will be free to withdraw from the study at any time.

Participants will not be paid for taking part in this study to avoid any possible feelings of coercion. However, in recompense for travel expenses, loss of earnings and the significant burden of repeated trips to the hospital, £10 for the screening visit and £500 for study visit 1, £350 for study visit 2,3, and 4, and £50 for study visit 5 will be made available for participants. This amount also reflects similar expense payments offered in previous studies by our research group.

5.7. **AUDITS AND INSPECTIONS**

The study may be subject to inspection and audit by Imperial College London under their remit as sponsor and other regulatory bodies to ensure adherence to GCP and The UK Policy Frame Work for Health and Social Care Research

6. **PUBLICATION POLICY**

The findings of the research will be published in an open-access, peer-reviewed journal. In addition we will be collaborating with patient groups and professional groups to disseminate the findings via multiple media channels such as patient association publications, print and broadcast media. No participants identifiable data will be included.

7. **REFERENCES**

Diabetes, U. K. (2014). Diabetes: facts and stats. *Diabetes UK*, *3*, 1-21.Nestel, P., Cehun, M., & Chronopoulos, A. (2004). Effects of long-term consumption and single meals of chickpeas on plasma glucose, insulin, and triacylglycerol concentrations. *The American journal of clinical nutrition*, *79*(3), 390-395.

Nestel, P., Cehun, M., & Chronopoulos, A. (2004). Effects of long-term consumption and single meals of chickpeas on plasma glucose, insulin, and triacylglycerol concentrations. *The American journal of clinical nutrition*, *79*(3), 390-395.

Pittaway, J. K., Ahuja, K. D., Robertson, I. K., & Ball, M. J. (2007). Effects of a controlled diet supplemented with chickpeas on serum lipids, glucose tolerance, satiety and bowel function. *Journal of the American College of Nutrition*, *26*(4), 334-340.

Raben, A., Tagliabue, A., Christensen, N. J., Madsen, J., Holst, J. J., & Astrup, A. (1994). Resistant starch: the effect on postprandial glycemia, hormonal response, and satiety. *The American journal of clinical nutrition*, *60*(4), 544-551.

Sensoy, I. (2014). A review on the relationship between food structure, processing, and bioavailability. *Critical reviews in food science and nutrition*, *54*(7), 902-909.

Smeets, A. J., Soenen, S., Luscombe-Marsh, N. D., Ueland, & Westerterp-Plantenga, M. S. (2008). Energy expenditure, satiety, and plasma ghrelin, glucagon-like peptide 1, and peptide tyrosine-tyrosine concentrations following a single high-protein lunch. *The Journal of nutrition*, *138*(4), 698-702.

Wang, Y. C., McPherson, K., Marsh, T., Gortmaker, S. L., & Brown, M. (2011). Health and economic burden of the projected obesity trends in the USA and the UK. *The Lancet, 378(9793), 815-825.*

Zafar, T. A., & Kabir, Y. (2017). Chickpeas suppress postprandial blood glucose concentration, and appetite and reduce energy intake at the next meal. *Journal of food science and technology*, *54*(4), 987-994.

Zhou, J., Martin, R. J., Tulley, R. T., Raggio, A. M., McCutcheon, K. L., Shen, L., ... & Keenan, M. J. (2008). Dietary resistant starch upregulates total GLP-1 and PYY in a sustained day-long manner through fermentation in rodents. *American Journal of Physiology-Endocrinology and Metabolism*, *295*(5), E1160-E1166.
